# Analysis of Clinicopathological Histomorphological and Molecular Differences in Right and Left Sided Colonic Carcinoma

**DOI:** 10.64898/2026.03.03.26347325

**Authors:** A.V. Tharangi Damayanthi

**Affiliations:** A retrospective study with an analytical component was done at the Department of Histopathology, National Hospital of Sri Lanka, Colombo

## Abstract

**Background:** Colorectal carcinoma (CRC) remains a significant cause of cancer morbidity and mortality worldwide. Right– and left-sided tumours differ in clinical, morphological, and molecular features. Microsatellite instability-high (MSI-H) tumours, often right-sided, are associated with distinct histopathological characteristics and prognostic implications. In Sri Lanka, molecular MSI testing is currently unavailable, highlighting the need for alternative predictive approaches.

**Objectives:** General Objective: To describe the clinical and histopathological characteristics of right– and left-sided colorectal cancers in a Sri Lankan cohort and evaluate their usefulness in predicting MSI-H tumours.

**Specific Objectives:** To compare clinicopathological features between right– and left-sided colorectal cancers.

To predict MSI-H tumours based on clinicopathological features, including assessment of the MsPath score and histological parameters.

To determine interobserver agreement for MsPath score application in selecting cases for MSI assessment.

**Methods:** A retrospective analytical study was conducted on 156 colorectal carcinoma resections diagnosed between 2019 and 2021 at the National Hospital of Sri Lanka. Histopathological evaluation included tumour differentiation, mucinous and medullary features, tumour-infiltrating lymphocytes (TILs), and Crohn-like reaction. MsPath scores were calculated based on age, tumour site, and histological parameters. Immunohistochemistry (IHC) for PMS2 and MSH6 was performed on 46 selected cases to assess mismatch repair (MMR) status.

**Results:** Of 156 cases, 41 (26%) were right-sided and 115 (74%) left-sided. The majority were moderately differentiated adenocarcinomas (89%). Histological features suggestive of MSI-H including TILs (29%) and Crohn-like lymphoid reaction (23%) were more frequent in right-sided tumours. MsPath scores ranged from 0.0 to 5.9, with 50% of cases scoring below 1. Among the 46 cases evaluated by IHC, MMR deficiency was observed predominantly in higher MsPath score categories, and a significant association was found between MsPath score category and MMR status (χ² = 13.76, df = 2, p = 0.001). Interobserver agreement for MsPath scoring was substantial (Kappa = xx, indicating reproducibility).

**Conclusion:** Right-sided colorectal carcinomas in this Sri Lankan cohort more frequently exhibited histological features suggestive of MSI-H, including mucinous differentiation, TILs, and Crohn-like lymphoid reaction. MsPath scoring correlated with MMR deficiency in the IHC-tested subset, but its predictive value is limited without immunohistochemical confirmation. IHC using a two-antibody panel (PMS2 and MSH6) proved to be a feasible, cost-effective, and reliable method for MSI screening in resource-limited settings. This study is the first comprehensive evaluation of right-versus left-sided colorectal carcinomas and MsPath utility in Sri Lanka, underscoring the need for expanded IHC capacity, larger cohorts, and integration of molecular testing for MSI validation.

## CHAPTER 1

### 1.1 Introduction

Out of the all cancer incidence in the world, colorectal carcinoma is the third commonest cancer (1). In 2018 in Sri Lanka, 1441 patients were diagnosed with colorectal carcinoma of which 734 were males and 707 were females. Incidence and mortality rates were 5.2 and 3.4(1)

The entire colon is about 150cm in length and consists of the caecum, ascending colon, hepatic flexure, transverse colon, splenic flexure, descending colon, sigmoid colon and rectum(2). Right colonic tumours include the tumours proximal to the splenic flexure.

Tumours in the right and left colon exhibit different molecular and histopathological features (2). Tumours with microsatellite instability-high (MSI-H) and CpG island methylation microsatellite stable (MMS) tumours are frequently located in the caecum, ascending colon and transverse colon. (3) Nearly 15% of sporadic colorectal cancers (CRC) and almost all CRCs in hereditary non polyposis colorectal carcinoma (HNPCC)/Lynch syndrome patients develop along the MSI pathway.

Microsatellites are nucleotide repeat sequences. They form insertion-deletion loops in DNA replication. Mismatch repair genes are MLH1, MSH2, MSH6 and PMS2 which help to keep microsatellite at germline length. Inactivation of one of the above four genes can cause microsatellite instability (MSI). Microdissection and polymerase chain reaction (PCR)-based detection are used to determine the MSI status of a tumour (4).

Right-sided tumours with mucinous features, poor differentiation (solid or medullary appearance), tumour infiltrating lymphocytes, lack of dirty necrosis and presence of a Crohn like inflammatory reaction are more likely to be MSI-H (4,6,7,8,9,13,14,15).

Detection of microsatellite instability in CRC is necessary to identify HNPCC as well as for prognostication and for decisions of treatment. However, tests to detect MSI are expensive and not widely available in Sri Lanka. So we are planning to conduct this study in two phases; in the first phase, to study the morphological features of right and left sided CRC and in the second phase, based on findings of the 1st phase, to select the carcinomas that may show a MSI-H and subject those tumours to molecular/ immunohistochemical testing with the funding available.

### 1.2 Objectives

#### General objective

To describe the clinical and histopathological characteristics of right sided and left sided colorectal tumours and to predict the tumours that may show high microsatellite instability.

#### Specific objectives

1. To describe the differences in clinicopathological features of right and left colonic tumours in a Sri Lankan setting
2. To predict the MSI-High colorectal cancers based on the clinicopathological features
3. To determine the interobserver agreement for the use of MsPath score in selecting cases for testing MSI status

### 1.3 Literature review

Chandrasinghe et al analyzed 679 patients, and identified significant increase of both colon and rectal cancers over time. The majority were left-sided cancers. They have also observed left-sided preponderance, younger age at presentation and advanced stage at presentation. They observed patterns of CRC are different in South Asian countries and in the western countries. This would have implications on treatment as well as in disease surveillance. (10)

As stated in different cohort studies, right-sided CRC commonly occurs in women and in elderly people while occurrence of left-sided CRC is mainly identified in younger men.

According to Burcin et al the following contrasts are noted between right-sided and left-sided CRC (06)

**Table 01.**
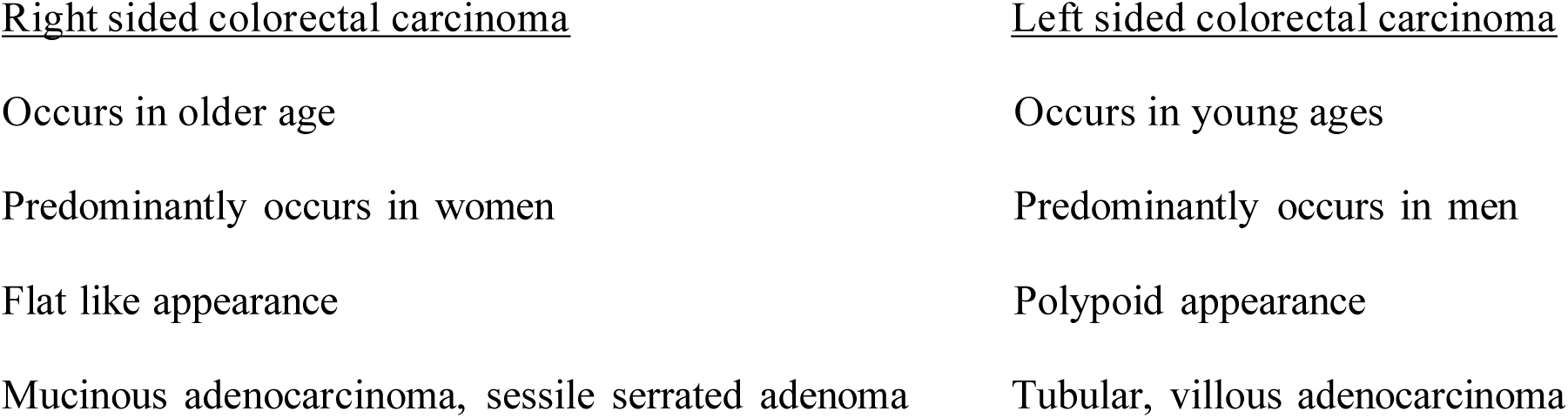
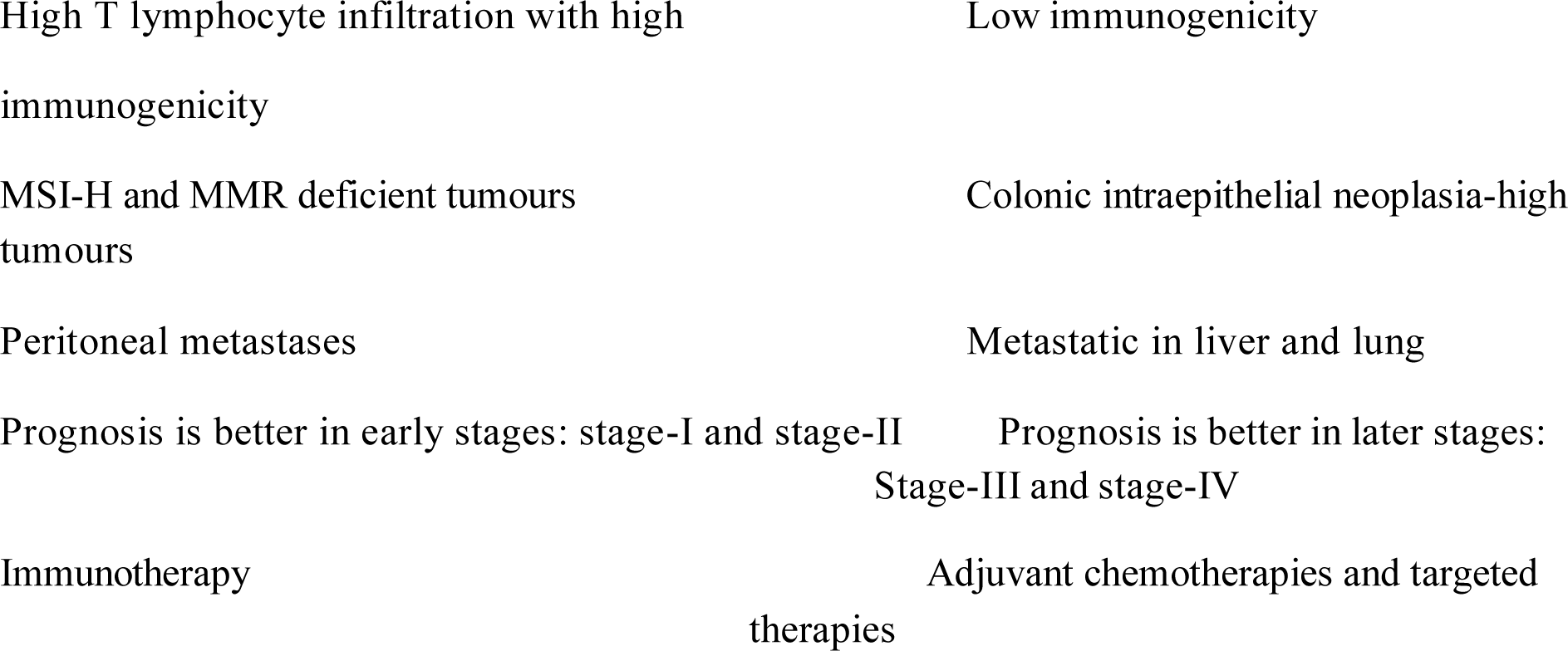
– Differences between right and left sided CRC.

**Table 02.**
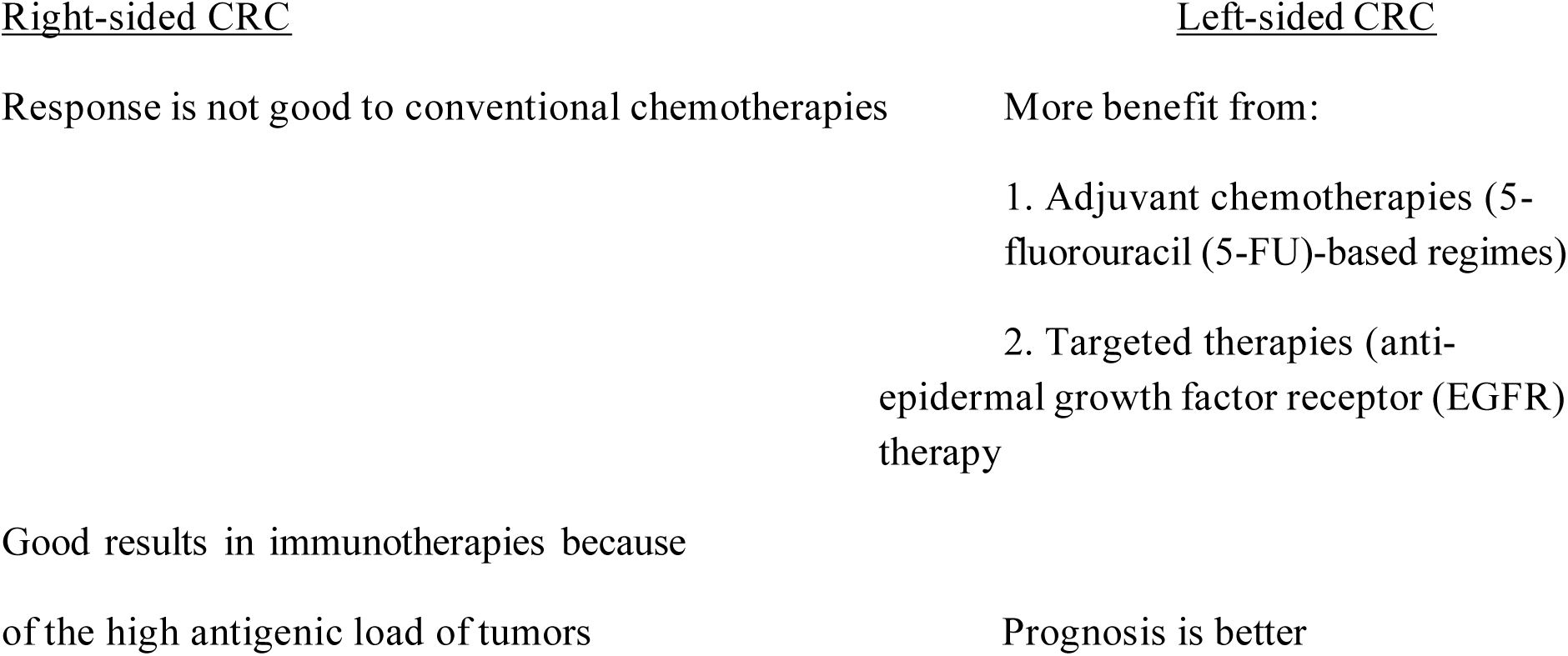
– Differences between treatment responses of the right and left CRC (06)

To develop efficacious treatments, it is required to consider left-sided tumors and right-sided tumors as two different entities, and consider the differences in designing the therapy regimes (6).

According to WHO classification of tumours of the digestive system (3) tumours with MSI-H, whether sporadic or Lynch associated, have morphological differences from tumours with chromosomal instability.

They are frequently right sided, mucinous or occasionally medullary type, associated with a Crohn like peritumoral infiltrate, intratumoral lymphocyte infiltrate, devoid of “dirty” necrosis, lower stage, expansile growth and better stage specific prognosis.

As mentioned in numerous publications histological common features in MSI-H CRCs are poor differentiation, mucinous type, right sided, lack of dirty necrosis, increased Tumour Infiltrating Lymphocytes(TIL), a circumscribed or expansile growth pattern, histologic heterogeneity and a prominent inflammatory reaction at the advancing edge of the tumour /Crohn like reaction (6,7,8,9,14,22,24,27).

Jenkins et al introduced a MsPath ((Microsatellite instability by Pathology) score after analyzing the histological features of patients under the age of 60,tumour-infiltrating lymphocyte, proximal subsite, mucinous type, Crohn-like reaction, poor differentiation and age <50 years(28).

Alexander et al mentioned that medullary-type carcinoma showed high specificity for MSI-H status and low sensitivity (7).

Smyrk et al studied on tumour infiltrating lymphocyte (TIL) counts and reported that TIL could reduce the number of CRCs referred for MSI testing by more than one-half, and still 93% of the MSI-high carcinomas would be identified (8).

Greenson et al did a similar study as Jenkins et al, on patients of all ages and found that right-sided location, >2 TIL per high-power field, lack of dirty necrosis, presence of a Crohn like reaction, any mucinous differentiation and poor differentiation, and <50 years were all independent predictors of MSI-H (9).

There are several methods to detect Microsatellite instability (33).

**Table 03.**
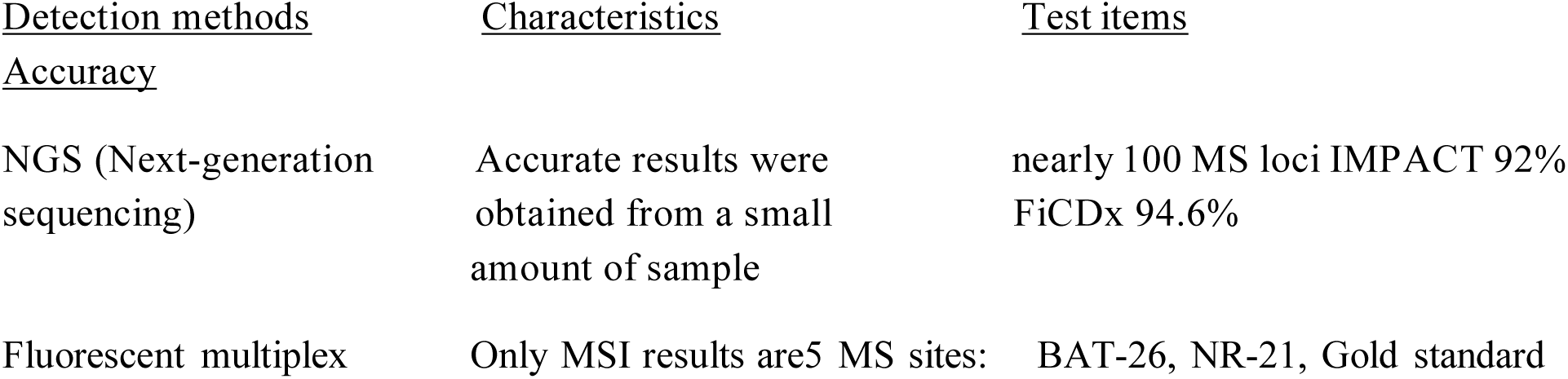
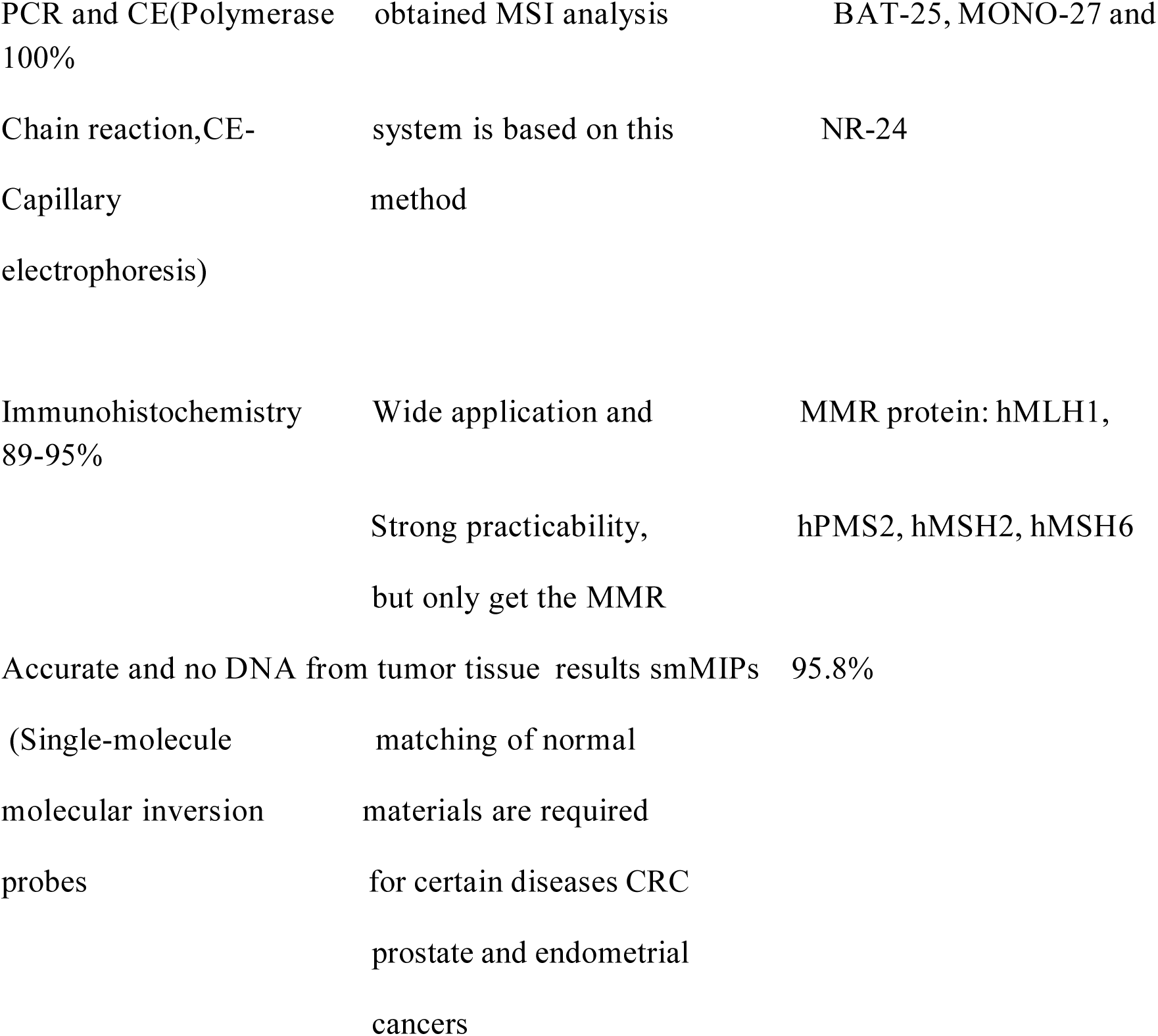
– Summary of microsatellite instability detection methods.

| <u>Detection methods</u> | <u>Characteristics</u> | <u>Test items</u> |
| --- | --- | --- |
| <u>Accuracy</u> |  |  |
| NGS (Next-generation sequencing) | Accurate results were obtained from a small amount of sample | nearly 100 MS loci IMPACT 92% FiCDx 94.6% |
| Fluorescent multiplex | Only MSI results are 5 MS sites: | BAT-26, NR-21, Gold standard |
| PCR and CE(Polymerase<br>100% | obtained MSI analysis | BAT-25, MONO-27 and |
| Chain reaction,CE-Capillary<br>electrophoresis) | system is based on this<br>method | NR-24 |
| Immunohistochemistry<br>89-95% | Wide application and<br>Strong practicability,<br>but only get the MMR | MMR protein: hMLH1,<br>hPMS2, hMSH2, hMSH6 |
| Accurate and no DNA from tumor tissue | results smMIPs | 95.8% |
| (Single-molecule<br>molecular inversion<br>probes | matching of normal<br>materials are required<br>for certain diseases CRC<br>prostate and endometrial<br>cancers |  |

Even though for immunohistochemical studies a 4-antibody panel that include the 4 most commonly affected proteins (MLH1, MSH2, MSH6, and PMS2) is being used generally, there are various studies describing the importance of two panel test comprising PMS2 and MSH6 (34,35).

Mojtahed et al described the effectiveness of two antibody panel including PMS2 and MSH6 by the action of mismatch repair proteins within a cell.

“MLH1 dimerizes with PMS2 and MSH2 dimerizes with MSH6. MLH1 and MSH2 are the obligatory partners for their respective heterodimers. In general, germline mutations in the obligatory partners MLH1 and MSH2 most often will result in degradation of the hetero dimer and the consequent proteolytic degradation of their respective secondary partners, PMS2 and MSH6. In contrast, germline mutations in the secondary partners MSH6 and PMS2 may not result in proteolytic degradation of its obligatory partner, as the function of the secondary protein may be compensated by other proteins.” So based on this concept they have done a study and concluded a two-antibody panel including PMS2 and MSH6 is a cost-effective screening approach for identifying MMR abnormalities in CRC, in patients suspected of having Lynch syndrome. Also their analysis indicates that the two-antibody panel approach is equally effective as the conventional four-antibody panel in screening for patients who harbor tumors with MMR protein abnormalities (34).

According to Hall et al the sensitivity and specificity of a two panel test comprising PMS2 and MSH6, compared to a four panel test, and is 100%. No false negatives or positives were identified. So they have concluded that the two panel test (PMS2 and MSH6) should replace a four panel protocol for immunohistochemical screening for mismatch repair deficiency (35).

### 1.4 Justification

This is the first large scale study to be conducted in Sri Lanka to compare the clinical and morphological features of tumours in the right and left colon. Also there are no previously reported studies done on the histopathological predictors of microsatellite instability in CRC in a Sri Lankan context.

It is beneficial to identify a certain CRC as MSI-H. Then there is a chance to detect Hereditary CRC prone syndromes, predict prognosis and potentially influence therapy.

The information gained from this study will be beneficial to assess whether these morphologic features should be added to the routine histopathology reports.

The findings of this study will contribute to the advancement of the general knowledge in the field of molecular testing and use of IHC to detect MSI-H colorectal carcinoma.

Also we hypothesize that insight gained from this correlative analysis will provide directions for future research efforts on colorectal carcinoma morphology as well as phenotype.

With the help of the above histological features, we hypothesize that; MSI-high status can be predicted by routine histological examinations. If the hypothesis is true, it can be used as fre e screen to find cases that require molecular assessment of MSI.

### 1.5 Study Materials

This was a retrospective study with an analytical component carried out in the Department of Histopathology at the National Hospital of Sri Lanka.

#### Sample Selection and Size

A total of 156 cases of colorectal carcinoma (CRC) assessed during the study period met the eligibility criteria and were included in the study.

#### Inclusion Criteria

Only resected specimens demonstrating invasive adenocarcinoma of the colon or rectum were included. In cases with synchronous carcinomas, the largest tumour was selected for assessment.

#### Exclusion Criteria

Cases were excluded if they had received neoadjuvant chemotherapy, showed only intramucosal carcinoma or carcinoma in situ, or had damaged or faded slides that could not be restored by re-staining.

#### Sample Size

For Specific Objective 1, all cases that fulfilled the inclusion criteria during the period from January 2019 to January 2021 were included, resulting in a total sample size of 156 cases.

### 1.6 Sampling Techniques, Histopathological Assessment, and MSPath Score Calculation

All haematoxylin and eosin (H&E) stained tumour-containing slides of each case were reviewed by the principal investigator without access to any clinical information to minimize observer bias. The following histopathological criteria were assessed according to the definitions described in the MSPath score.

#### 1. Poor differentiation

The degree of differentiation was determined based on the architectural pattern, specifically the extent of gland or tubule formation. Poor differentiation was defined as a tumour demonstrating at least some glandular structures and/or mucin production, but with glands that were highly irregular and difficult to discern. The grade of differentiation was assessed based on the least differentiated area of the tumour, excluding regions of dedifferentiation or tumour budding at the invasive margin. (28)

#### 2. Histological type

Mucinous adenocarcinoma: Classified as mucinous adenocarcinoma when >50% of the tumour consisted of pools of extracellular mucin containing malignant epithelial cells in clusters, layers, or as individual cells, including signet ring cells. (3, 7, 9, 28)

Signet ring cell carcinoma: Defined by the presence of >50% tumour cells exhibiting prominent intracytoplasmic mucin with characteristic displacement or moulding of the nucleus. (3, 7, 9, 28)

Medullary carcinoma: Characterized by solid sheets of malignant cells with vesicular nuclei, prominent nucleoli, and abundant eosinophilic cytoplasm, accompanied by a prominent lymphocytic and neutrophilic infiltrate. (3, 28)

Undifferentiated carcinoma: Tumours that lack morphological evidence of differentiation beyond that of an epithelial tumour. (3)

#### 3. Tumour-infiltrating lymphocytes (TILs)

TILs were recorded as present when at least five intraepithelial lymphocytes were identified in at least one high-power field (×40) after examining a minimum of ten high-power fields. (28)

#### 4. Crohn –like lymphocytic reaction

This feature was considered present when four or more nodular lymphoid aggregates were identified in a low-power field (×4) beyond the advancing edge of the tumour, typically within the subserosal or mesenteric fat. (28)

The histological features were assessed while the observers were blinded to the tumour site. All collected information was documented in a data collection sheet (Annexure 1). No personal identification details were included in the data collection sheet. Each sample was identified using a unique research serial number to prevent direct patient identification. Identification details were accessible only to the principal investigator and were password-protected. After assessing the histological features, clinical information such as age, gender, and tumour site was obtained from the request form.

##### Calculation of MSPath score

For each case, the MSPath score was calculated by summing the weighted scores of individual clinicopathological and histopathological features, according to the coefficients established in the original scoring system. Each feature was scored as present (1) or absent (0), multiplied by its respective coefficient, and then summed to generate the total score (Annexure 2). The MSPath score was subsequently used to stratify tumours for further analysis, including selection for immunohistochemistry (IHC).

**Table 04.**
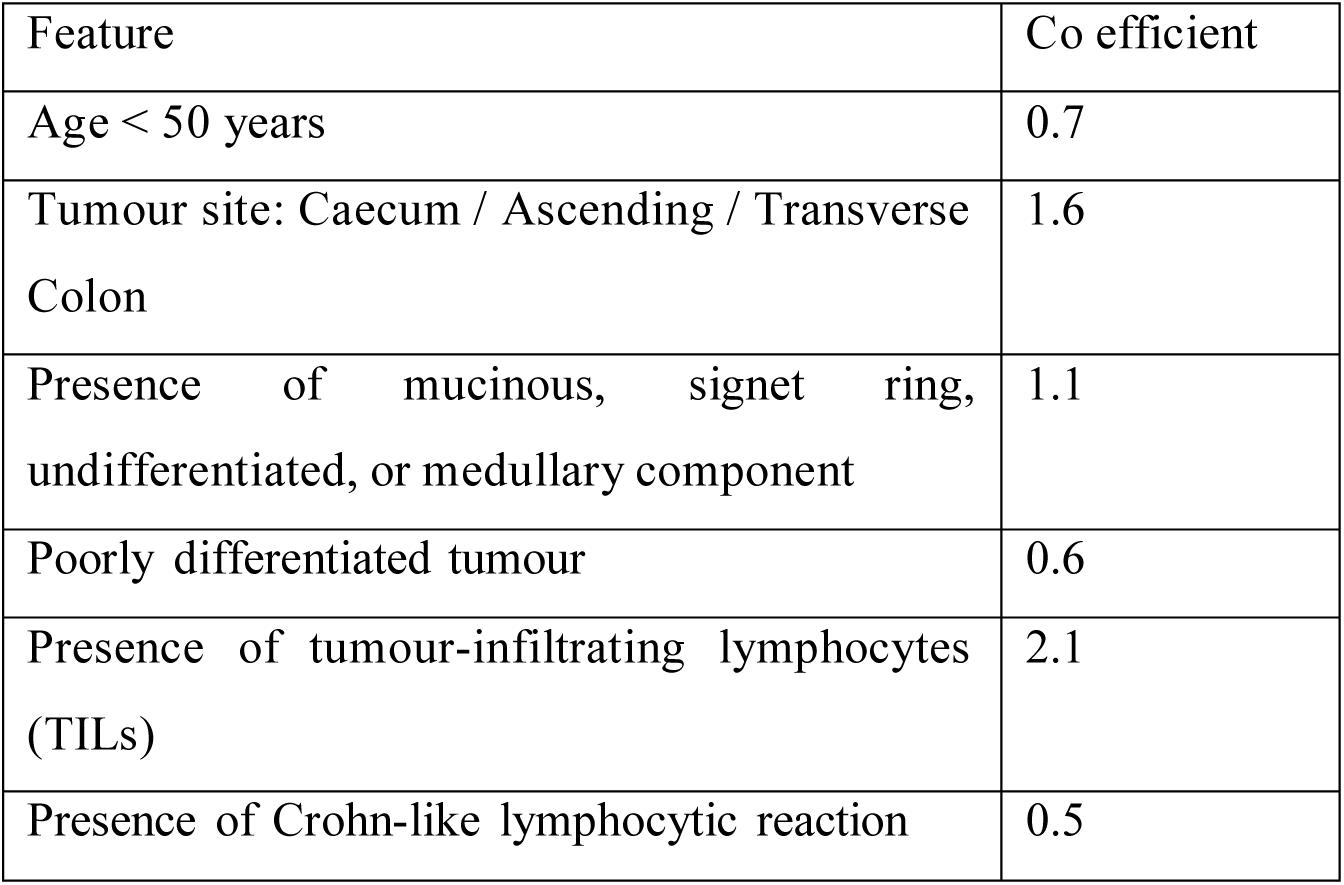
– MsPath score coefficients.

Tumours were categorized as MSPath score above or below the cut-off value of 1, in accordance with the original scoring system, to guide subsequent IHC analysis and correlate clinicopathological features with mismatch repair (MMR) status.

### 1.7 Inter-observer Agreement

To evaluate the inter-observer agreement in the assessment of MSPath score, a random sample of 30% of cases above and below the MSPath score cut-off of 1 was selected by the principal investigator (46 cases). An independent histopathology trainee of equivalent level was involved in the review process.

The trainee was provided with the same set of H&E-stained slides for the selected cases and the structured datasheet including definitions for each histological criterion relevant to the MSPath score.

The trainee independently assessed the individual histological features and calculated the total MSPath score for each case. In this study, no differences were observed between the principal investigator and the trainee, as the predefined selection criteria for histological features and MSPath score were highly specific and unambiguous. This confirmed excellent inter-observer agreement for both individual features and total MSPath scores.

Wax blocks for the selected 46 cases had been traced.

A two-panel immunohistochemistry (IHC) test, comprising PMS2 and MSH6, was performed (Annexure 3). Appendix tissue and endometrial tissue were used as positive controls for PMS2 and MSH6, respectively. All procedures were carried out at the Immunohistochemistry Laboratory, Medical Research Institute, Colombo.

### 1.8 Funding and selection for immunohistochemistry (IHC)

Funds were obtained from the Medical Research Institute (MRI) to perform immunohistochemical (IHC) analysis. Initially, IHC was planned to be carried out on approximately 25% of the total study cases. However, due to the availability of additional funding, 46 cases (approximately 30%) were included for IHC evaluation.

### 1.9 Immunohistochemistry Assessments and Interpretation

All IHC-stained slides were examined using a light microscope. Tumours were categorized as positive (antigen detected) when >10% of malignant cells demonstrated nuclear staining. Tumours were recorded as negative (antigen not detected) if nuclear staining was absent in all malignant cells, while normal epithelial and stromal cells retained positive staining, serving as internal controls.

Any errors, pitfalls, and limitations encountered during the assessment of histological features and IHC were documented and described in detail to ensure transparency and reproducibility of the study.

### 1.10 Data analysis

Statistical comparisons between the categories were done using chi-square (X2 test). The level of statistical significance was regarded as < 0.05 for this study.

### 1.11 Ethical Clearance

The ethical clearance was obtained from the Ethics Review Committee, National Hospital of Sri Lanka.

## CHAPTER 02

### Results

#### 2.1 Demographic Characteristics

A total of 156 colorectal carcinoma (CRC) resection specimens were included in the study. The patients’ ages ranged from 31 to 90 years, with a median of 64 years.

All cases were reviewed for clinicopathological features. (n = 156) 11 cases (7%) showed the presence of two tumours within a single specimen. These combinations included:

**Figure 01.**
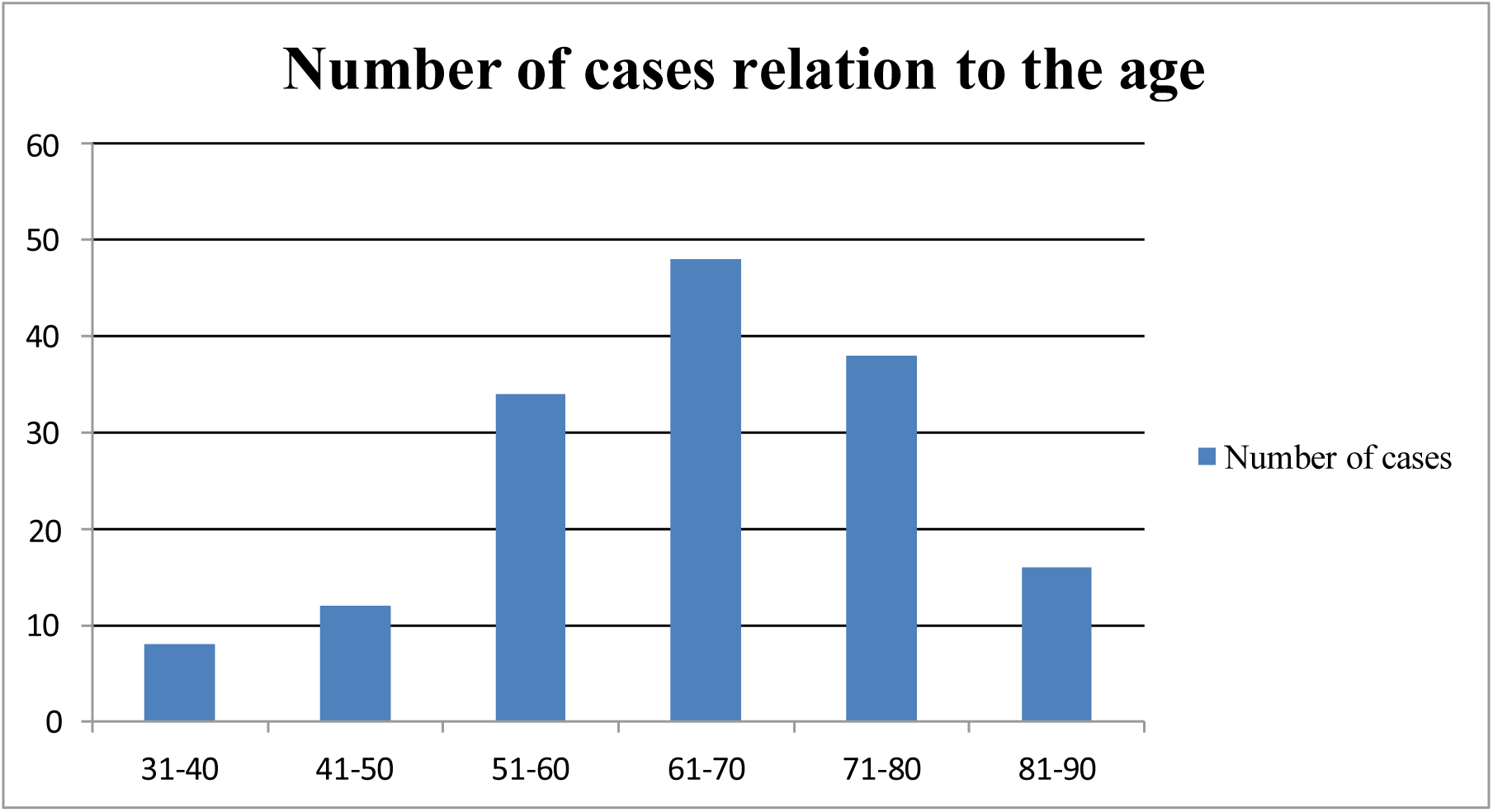

**Figure 02.**
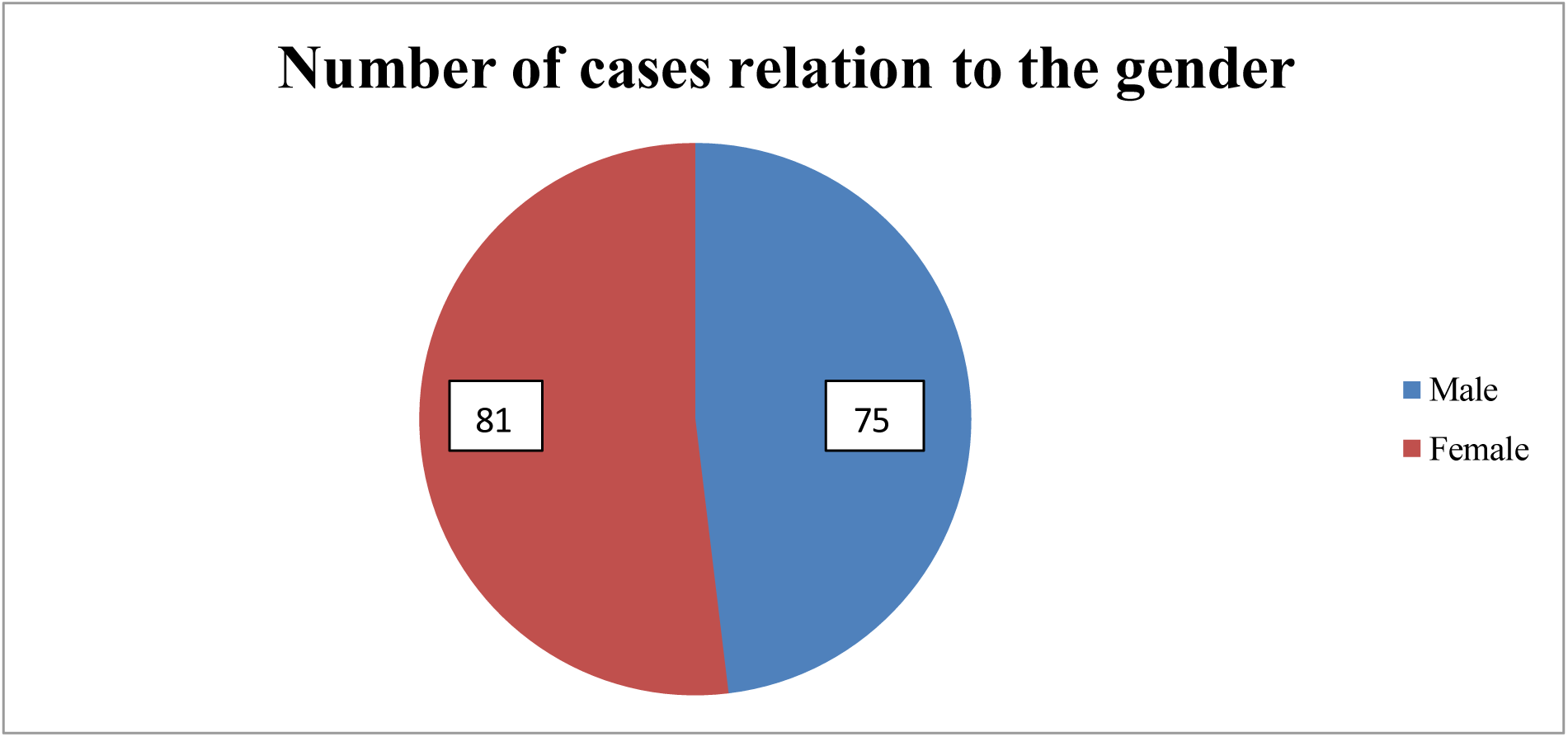

**Table 05.** – Demographic data.

| Age range | Number of cases in males | Number of cases in Females | Total Number of cases | Percentage |
| --- | --- | --- | --- | --- |
| 31-40 | 4 | 4 | 8 | 5% |
| 41-50 | 5 | 7 | 12 | 8% |
| 51-60 | 13 | 21 | 34 | 22% |
| 61-70 | 25 | 23 | 48 | 31% |
| 71-80 | 19 | 19 | 38 | 24% |
| 81-90 | 9 | 7 | 16 | 10% |

**Table 06.** – Case distribution by sex.

| Sex | Number of cases | Percentage |
| --- | --- | --- |
| Male | 75 | 48% |
| Female | 81 | 52% |

**Table 07.** – Distribution of Tumour Sites.

| Tumour Site | Number of Cases | Percentage (%) |
| --- | --- | --- |
| Caecum | 10 | 6.4% |
| Ascending colon | 7 | 4.8% |
| Transverse colon | 15 | 9.6% |
| Descending colon | 7 | 4.4% |
| Sigmoid colon | 57 | 36.5% |
| Rectosigmoid junction | 14 | 8.9% |
| Rectum | 35 | 22.4% |
| Multiple tumours | 11 | 7% |

Caecum and ascending colon (7 cases), Caecum and transverse colon (1case), Ascending colon and transverse colon (1 cases), Sigmoid colon and rectum (1 case),Sigmoid colon and descending colon (1 case).

**Figure 03.**
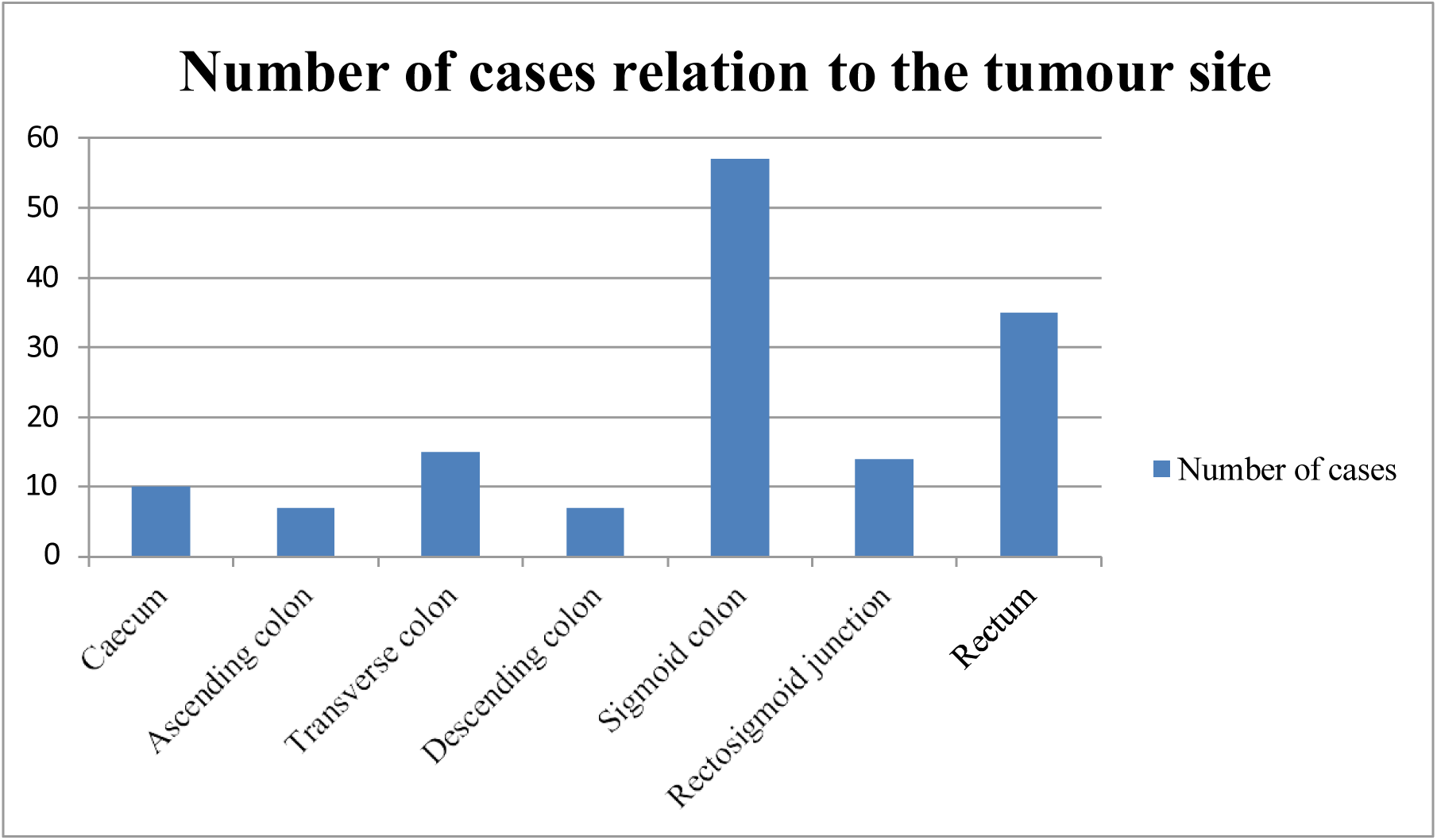

Right-sided colonic tumours were defined as those located proximal to the splenic flexure. Accordingly, tumours of the caecum, ascending colon and transverse colon were considered right-sided in this study. A total of 41 cases were classified as right sided, including nine cases with synchronous tumours: seven involving the caecum and ascending colon, one involving caecum and transverse colon and another one involving the ascending colon and transverse colon.

A total of 115 cases were classified as left-sided, including two cases with synchronous tumours: one involving the sigmoid colon and rectum, and another involving the sigmoid and descending colon.

**Figure 04.**
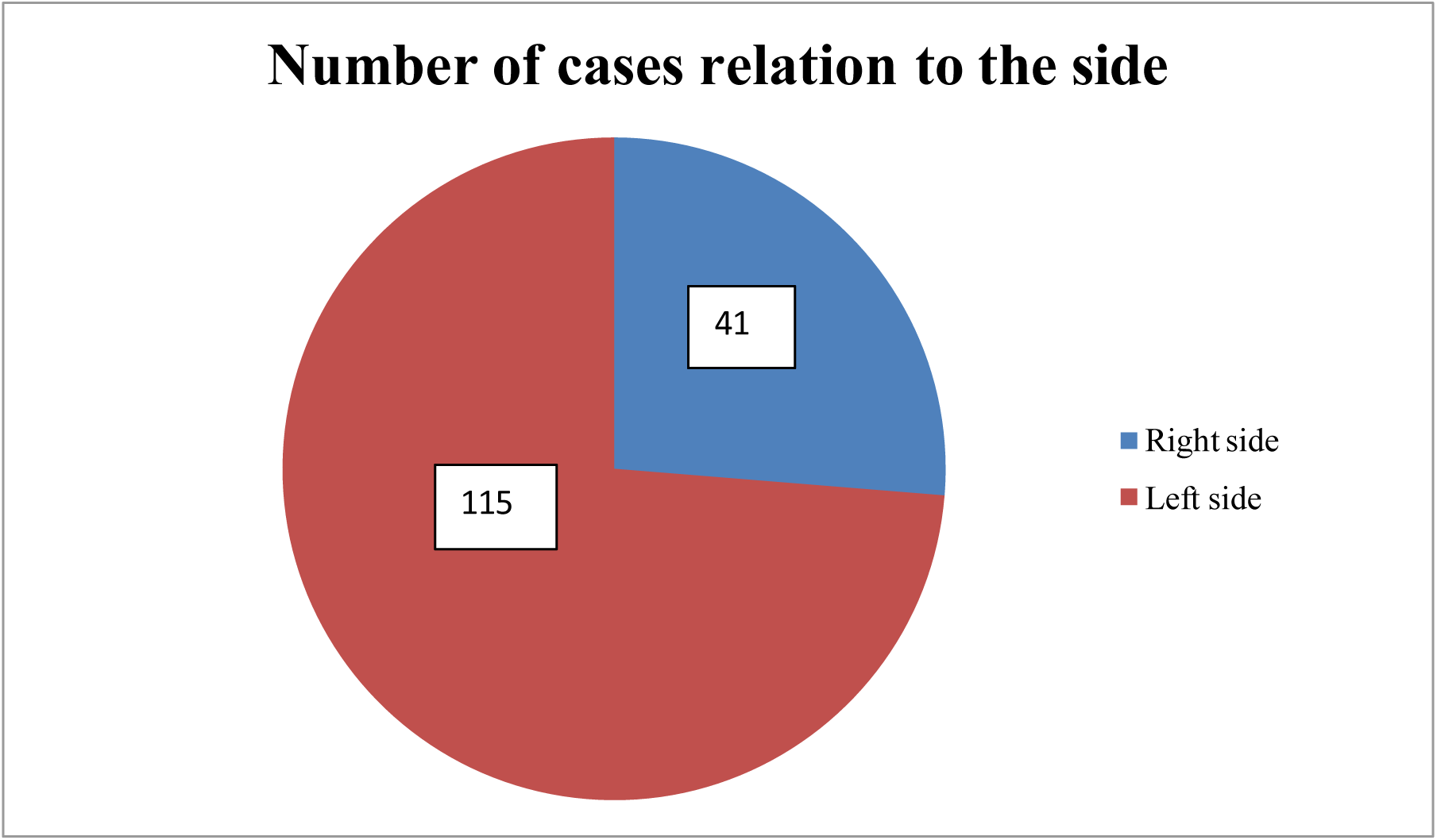

#### 2.2 Histopathological Characteristics

Regarding tumour differentiation, the vast majority of cases were moderately differentiated adenocarcinoma, NOS.

**Table 08.** – Distribution of tumour differentiation.

| Histological type | Number of cases | Percentage |
| --- | --- | --- |
| Low grade (moderately differentiated adenocarcinoma, NOS) | 139 | 89.1% |
| High grade (Poorly differentiated adenocarcinoma, NOS) | 1 | 0.6% |
| Adenocarcinoma NOS with mucinous component | 14 | 9% |
| Mucinous adenocarcinoma | 1 | 0.6% |
| Undifferentiated carcinoma | 1 | 0.6% |
| Signet ring cell component/carcinoma | 0 | 0 |
| Medullary carcinoma | 0 | 0 |

Poorly differentiated carcinoma was observed in a male patient involving the caecum, aged between 50–55 years.

Undifferentiated carcinoma was observed in a female patient involving the rectum, aged between 60–65 years.

Tumors with greater than 50% area showing extracellular mucin were classified as mucinous carcinoma. Tumors with less than 50% area showing extracellular mucin were classified as having adenocarcinoma, NOS with mucinous component.

71% of adenocarcinoma with mucinous component is seen in left side of the colon.

**Figure 05.**
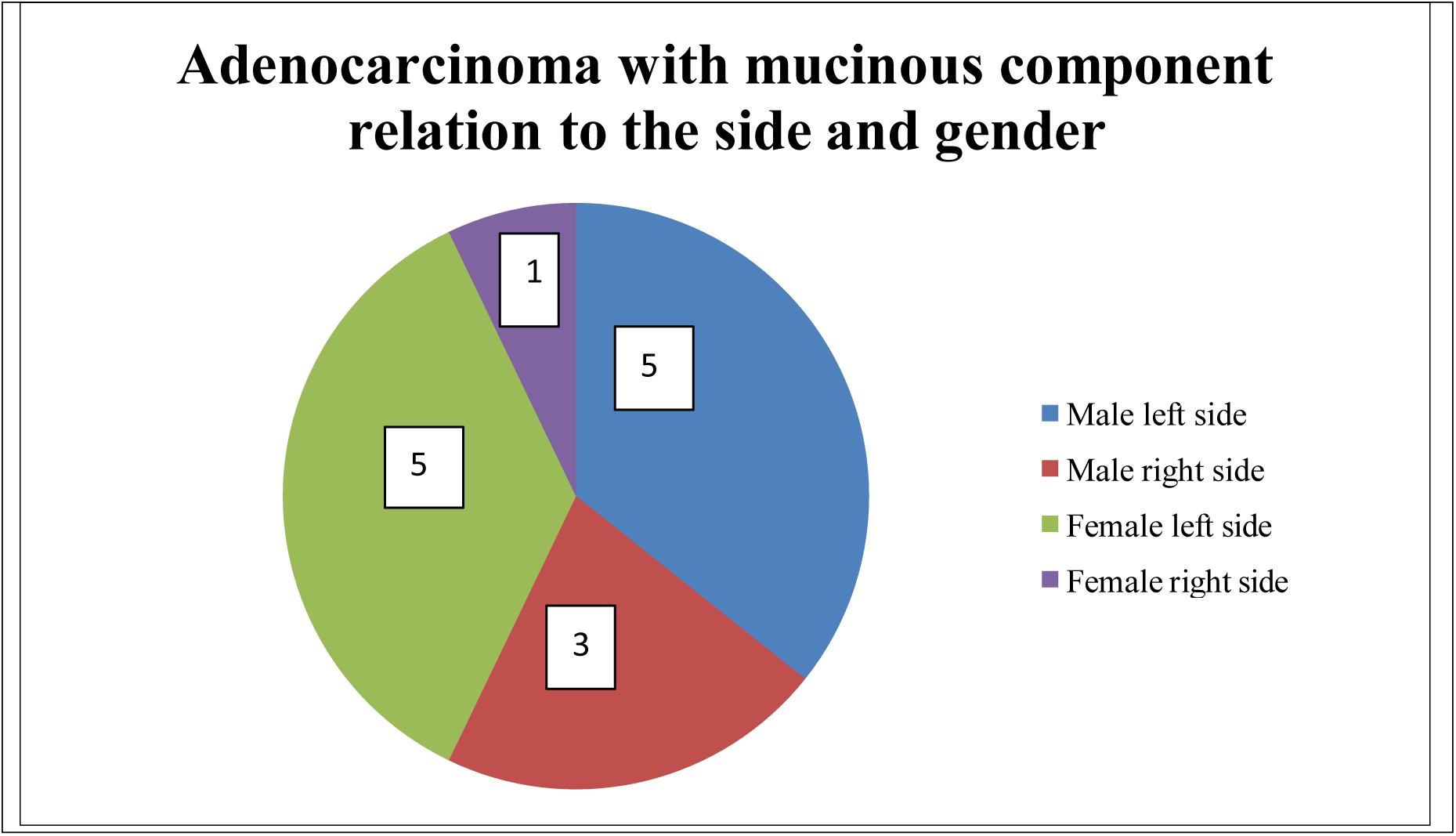

Immunological/Inflammatory Features

Tumour-infiltrating lymphocytes (TILs) were present in 45 cases (29%).

**Figure 06.**
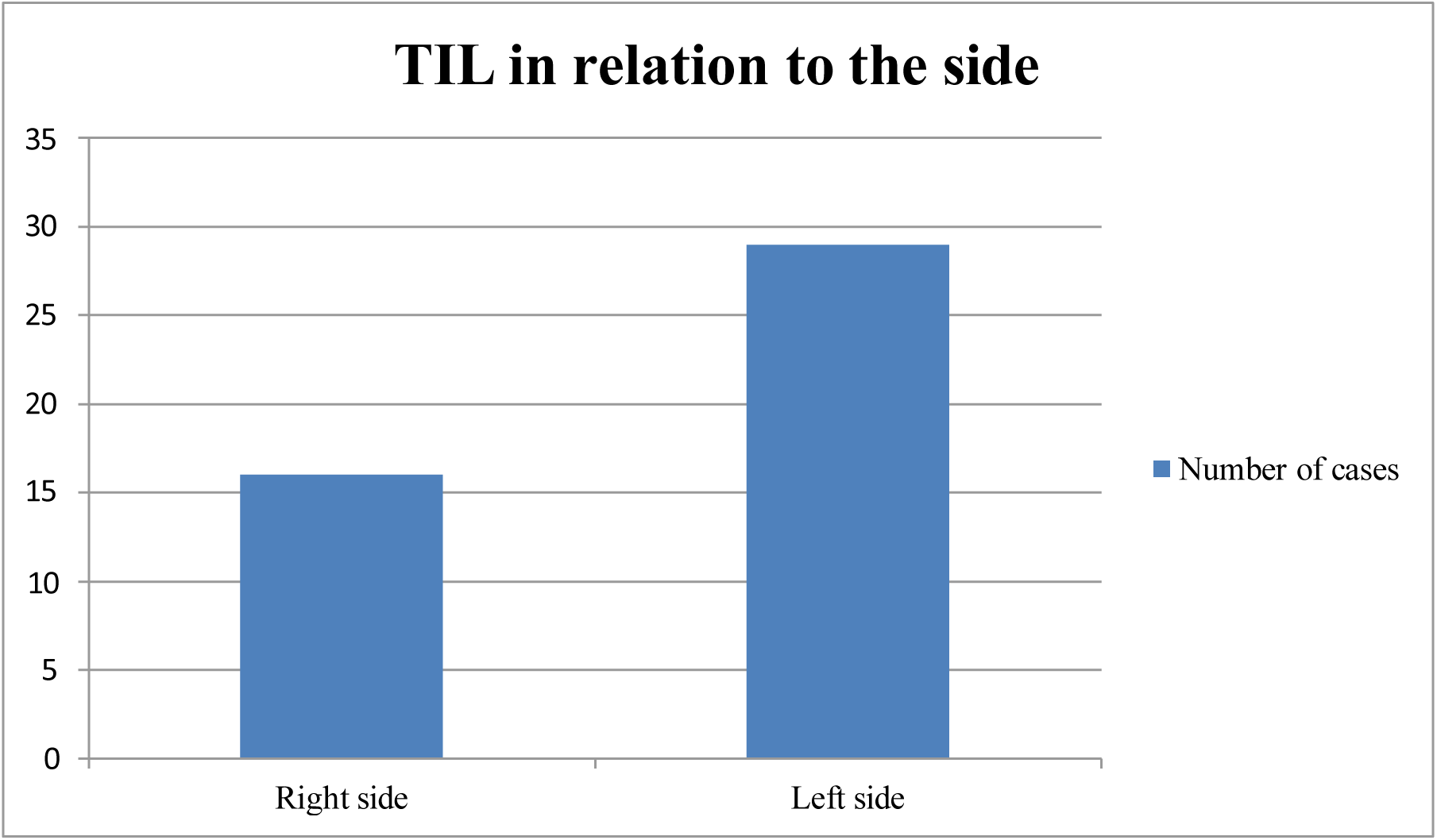

A Crohn-like lymphocytic reaction was identified in 36 cases (23%) Figure 07

**Figure 07.**
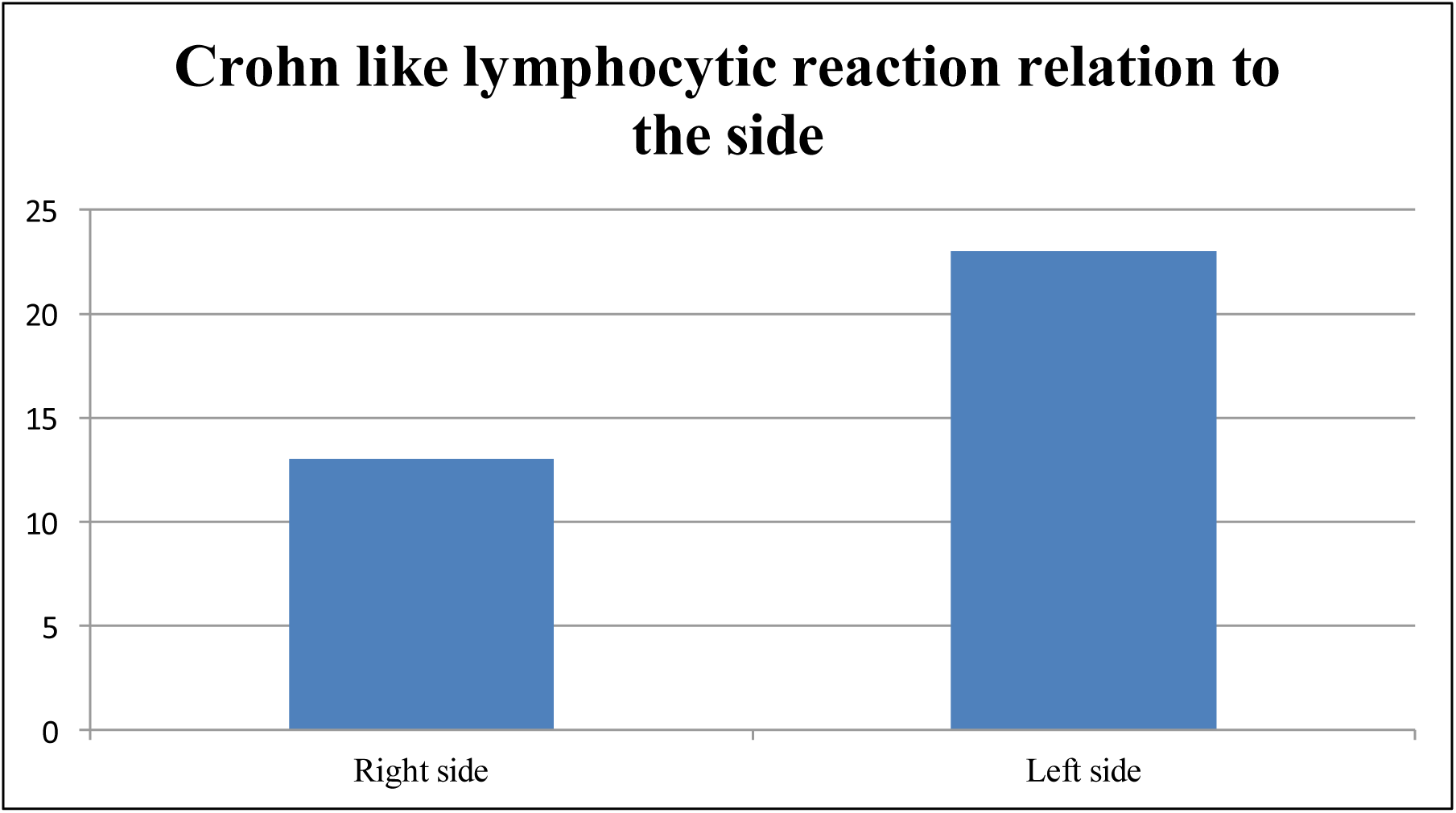

MsPath Score Distribution (n = 156)

This map shows the MSI probability score (sum of coefficients from table 4) along the top and the percent likelihood of microsatellite instability on the bottom. Hence a tumor with a MSI probability score of 0 would have a 1% chance of being MSI-H while a tumor with a MSI probability score of 4.5 would have a 50% chance of being MSI-H.

Paraffin-embedded tissue blocks from 30% of randomly selected colorectal carcinoma cases with sum scores above and below ‘1’ were evaluated for mismatch repair protein expression using a two-panel immunohistochemical test comprising PMS2 and MSH6.

**Table 09.**
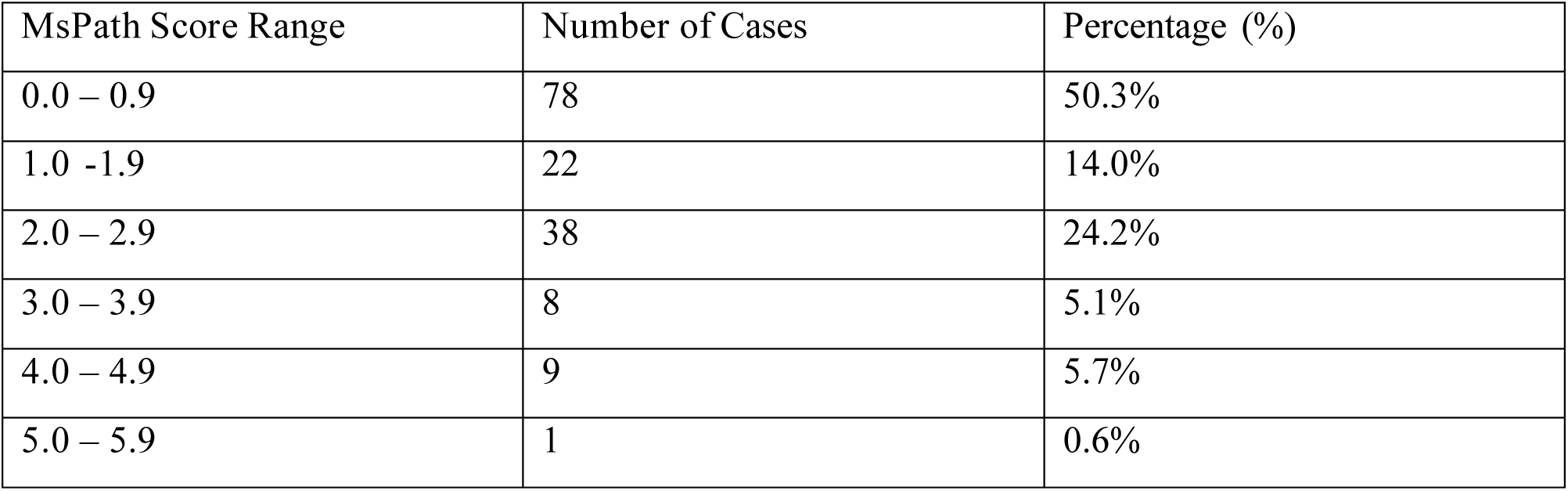
– MsPath score distribution.

78 cases show total MsPath score ≥1. Cut-off level of ‘1’ has been recommended for the MsPath score to maximize the specificity while maintaining a high sensitivity, because it is important not to miss MSI-H cases.(28)

**Table 10.**
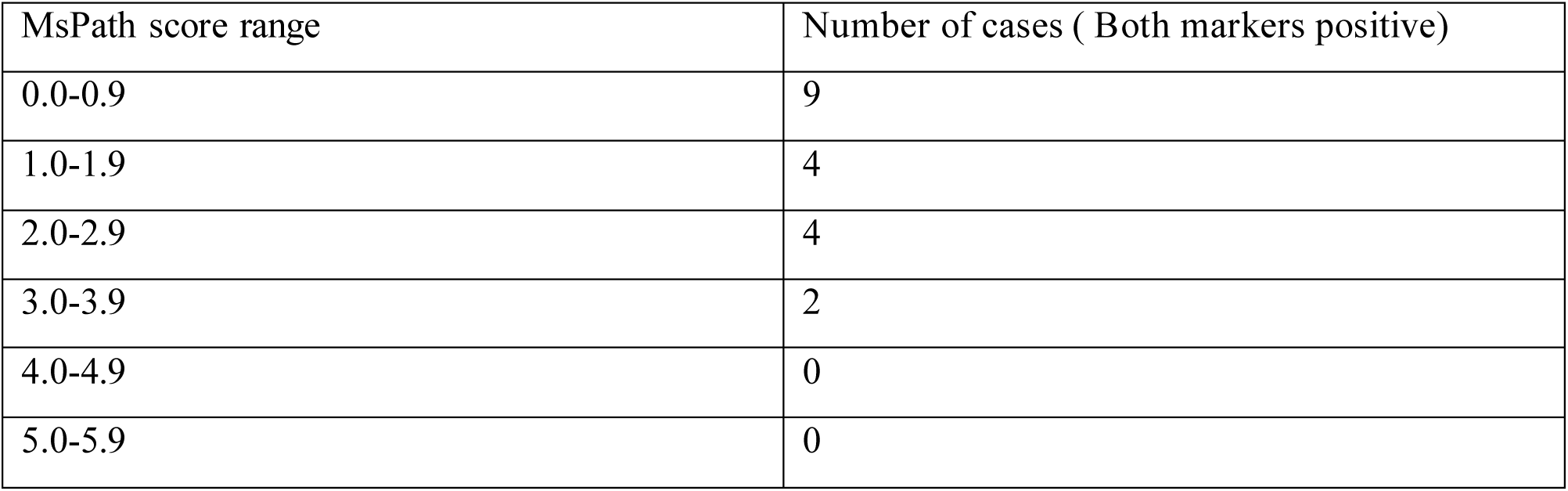
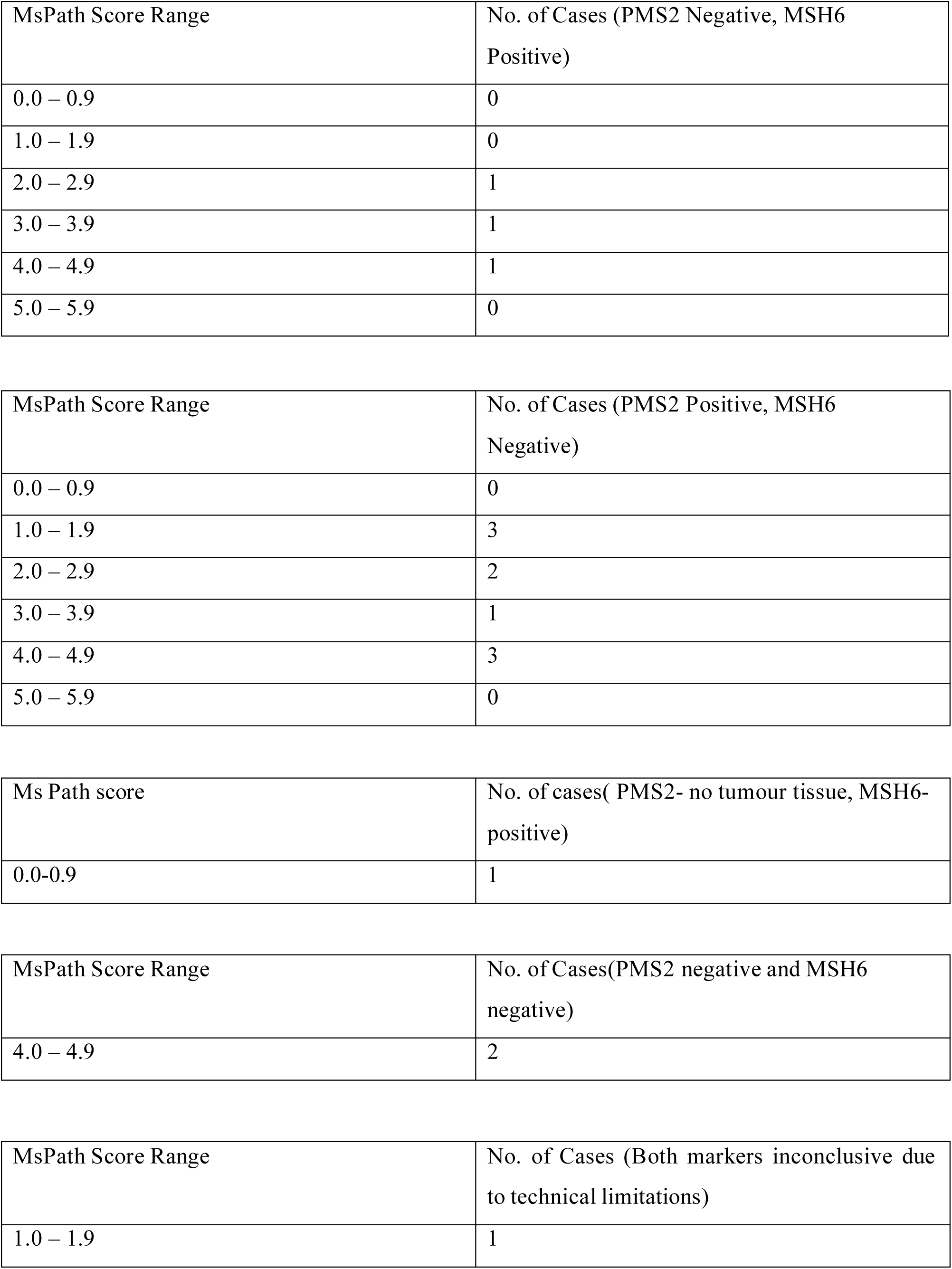
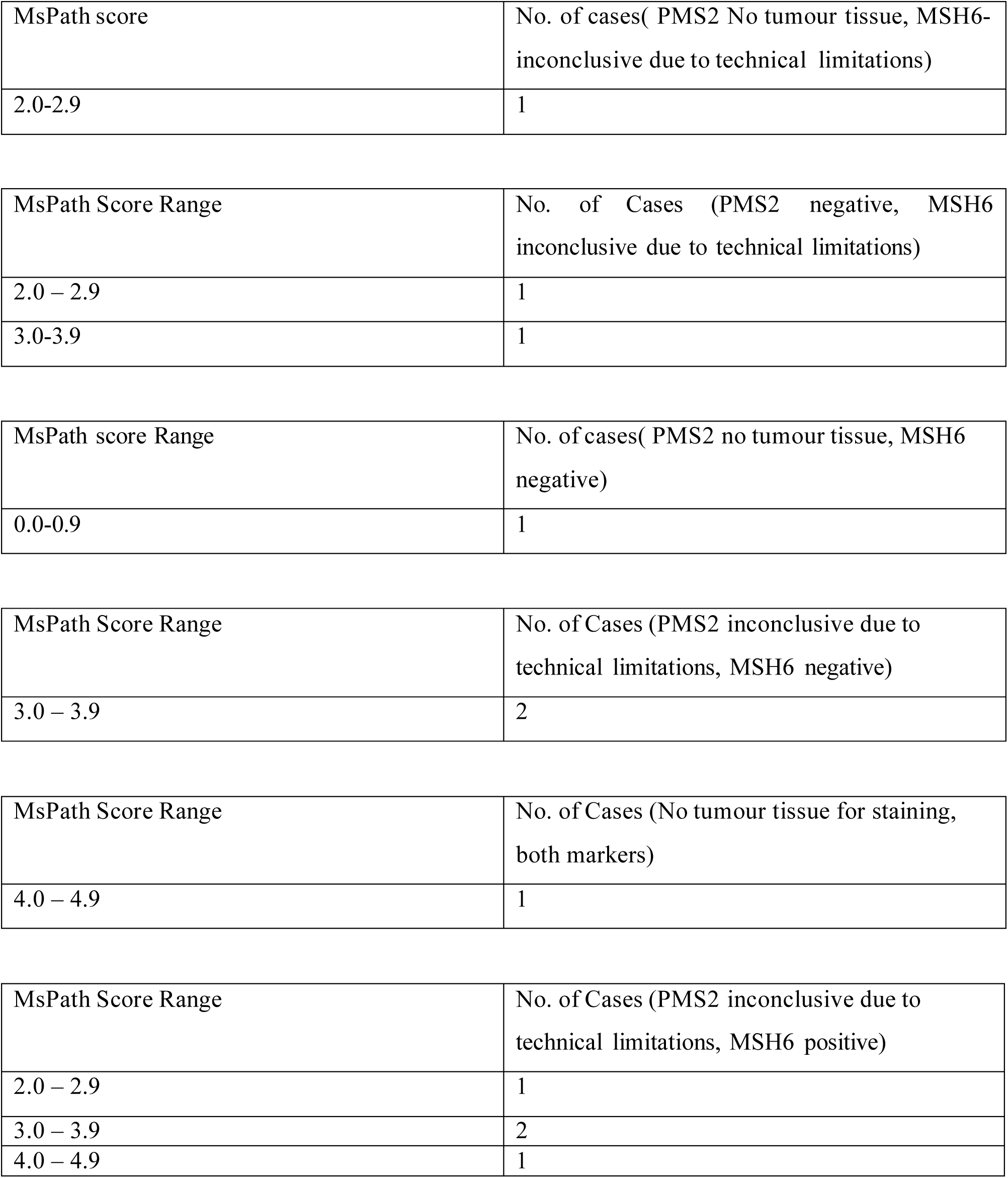
– The relationship of MsPath score and immunohistochemistry results.

| MsPath score range | Number of cases ( Both markers positive) |
| --- | --- |
| 0.0-0.9 | 9 |
| 1.0-1.9 | 4 |
| 2.0-2.9 | 4 |
| 3.0-3.9 | 2 |
| 4.0-4.9 | 0 |
| 5.0-5.9 | 0 |

| MsPath Score Range | No. of Cases (PMS2 Negative, MSH6 Positive) |
| --- | --- |
| 0.0 – 0.9 | 0 |
| 1.0 – 1.9 | 0 |
| 2.0 – 2.9 | 1 |
| 3.0 – 3.9 | 1 |
| 4.0 – 4.9 | 1 |
| 5.0 – 5.9 | 0 |

| MsPath Score Range | No. of Cases (PMS2 Positive, MSH6 Negative) |
| --- | --- |
| 0.0 – 0.9 | 0 |
| 1.0 – 1.9 | 3 |
| 2.0 – 2.9 | 2 |
| 3.0 – 3.9 | 1 |
| 4.0 – 4.9 | 3 |
| 5.0 – 5.9 | 0 |

| Ms Path score | No. of cases( PMS2- no tumour tissue, MSH6-positive) |
| --- | --- |
| 0.0-0.9 | 1 |

| MsPath Score Range | No. of Cases(PMS2 negative and MSH6 negative) |
| --- | --- |
| 4.0 – 4.9 | 2 |

| MsPath Score Range | No. of Cases (Both markers inconclusive due to technical limitations) |
| --- | --- |
| 1.0 – 1.9 | 1 |

| MsPath score | No. of cases( PMS2 No tumour tissue, MSH6-inconclusive due to technical limitations) |
| --- | --- |
| 2.0-2.9 | 1 |

| MsPath Score Range | No. of Cases (PMS2 negative, MSH6 inconclusive due to technical limitations) |
| --- | --- |
| 2.0 – 2.9 | 1 |
| 3.0-3.9 | 1 |

| MsPath score Range | No. of cases( PMS2 no tumour tissue, MSH6 negative) |
| --- | --- |
| 0.0-0.9 | 1 |

| MsPath Score Range | No. of Cases (PMS2 inconclusive due to technical limitations, MSH6 negative) |
| --- | --- |
| 3.0 – 3.9 | 2 |

| MsPath Score Range | No. of Cases (No tumour tissue for staining, both markers) |
| --- | --- |
| 4.0 – 4.9 | 1 |

| MsPath Score Range | No. of Cases (PMS2 inconclusive due to technical limitations, MSH6 positive) |
| --- | --- |
| 2.0 – 2.9 | 1 |
| 3.0 – 3.9 | 2 |
| 4.0 – 4.9 | 1 |

**Table 11.**
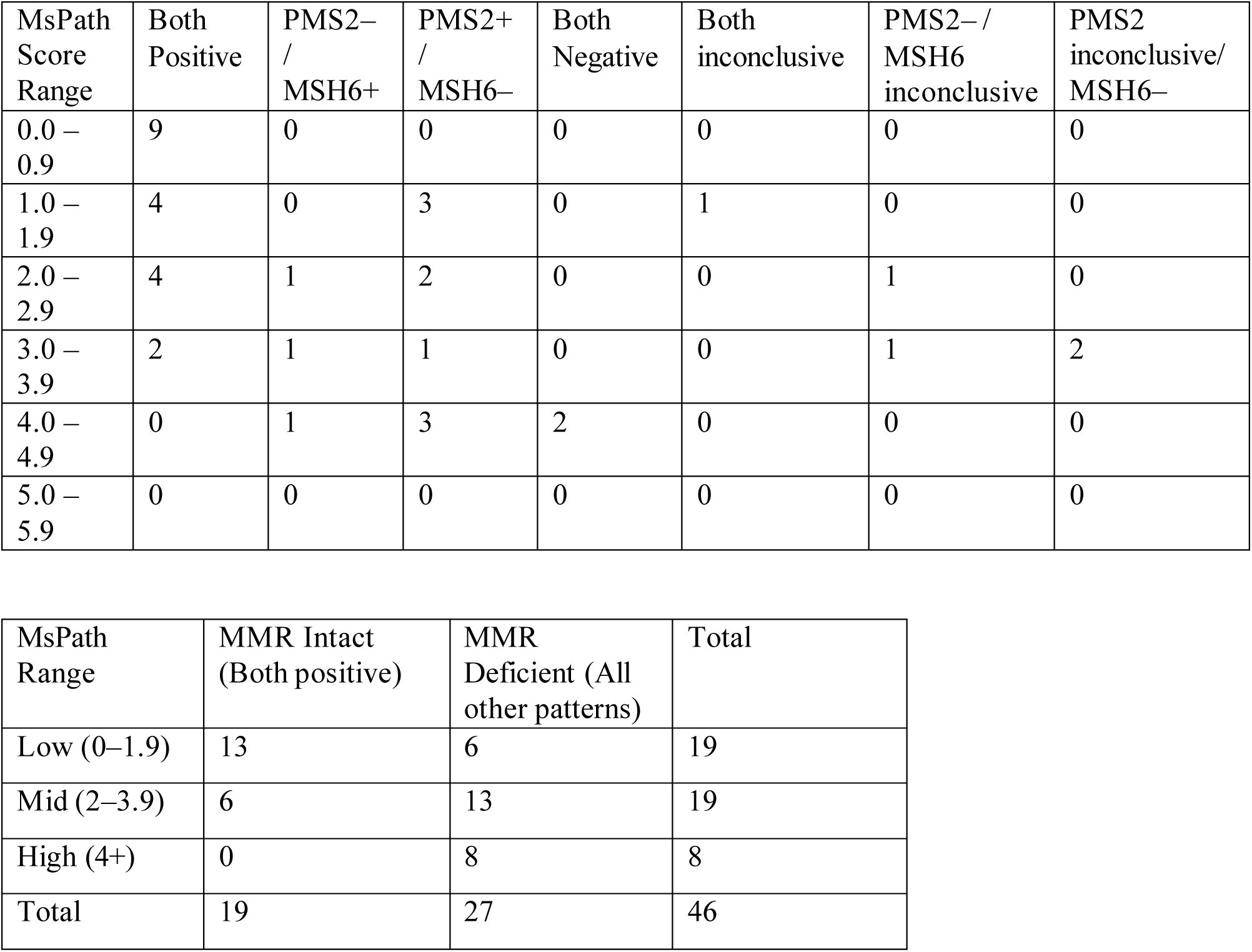
– Summary PMS2 and MSH6 IHC Status vs MsPath Score Ranges.

| MsPath Score Range | Both Positive | PMS2- / MSH6+ | PMS2+ / MSH6- | Both Negative | Both inconclusive | PMS2- / MSH6 inconclusive | PMS2 inconclusive/ MSH6- |
| --- | --- | --- | --- | --- | --- | --- | --- |
| 0.0 – 0.9 | 9 | 0 | 0 | 0 | 0 | 0 | 0 |
| 1.0 – 1.9 | 4 | 0 | 3 | 0 | 1 | 0 | 0 |
| 2.0 – 2.9 | 4 | 1 | 2 | 0 | 0 | 1 | 0 |
| 3.0 – 3.9 | 2 | 1 | 1 | 0 | 0 | 1 | 2 |
| 4.0 – 4.9 | 0 | 1 | 3 | 2 | 0 | 0 | 0 |
| 5.0 – 5.9 | 0 | 0 | 0 | 0 | 0 | 0 | 0 |

| MsPath Range | MMR Intact (Both positive) | MMR Deficient (All other patterns) | Total |
| --- | --- | --- | --- |
| Low (0–1.9) | 13 | 6 | 19 |
| Mid (2–3.9) | 6 | 13 | 19 |
| High (4+) | 0 | 8 | 8 |
| Total | 19 | 27 | 46 |

#### 2.3 Stastical analysis

Objective 2: Prediction of MSI-High colorectal cancers using clinicopathological features

Descriptive Analysis

Clinicopathological features were assessed in colorectal cancers stratified by location (right vs left side) and MsPath score.

Qualitative variables (e.g.,mucinous differentiation, tumour-infiltrating lymphocytes, poorly differentiated clusters) were analyzed using Chi-square tests to compare prevalence in right– and left-sided tumours.

Quantitative variables (e.g., tumour size, age at diagnosis) were first assessed for normality using the Kolmogorov-Smirnov and Shapiro-Wilk tests.

Normally distributed variables were compared using pooled t-tests. Non-normally distributed variables were compared using Mann-Whitney U tests.MSI Prediction Using MsPath Score and Histological Features.

Sensitivity, specificity, positive predictive value (PPV), and negative predictive value (NPV) were calculated for the MsPath score and each histological feature in predicting MSI-H status.

Binary logistic regression was applied to identify independent predictors of MSI-H status. Features significantly associated with MSI-H in univariate analyses (p < 0.05) were included in the multivariate model.

<u>Key predictors identified included</u>:

- High MsPath score (≥4.0)
- Presence of mucinous differentiation
- Tumour-infiltrating lymphocytes

These findings suggest that certain clinicopathological features, along with MsPath score, can reliably predict MSI-H colorectal cancers.

Objective 3: Interobserver Agreement for MsPath Score

<u>Agreement Analysis</u>

MsPath scoring was independently performed by two observers.

Agreed proportion: Number and percentage of cases where both observers assigned the same MsPath category.

Disagreed proportion: Number and percentage of cases with differing scores, with reasons for disagreement categorized (e.g., borderline histological features, poor tissue preservation, interpretational differences).

Kappa Coefficient

A Kappa statistic was calculated to quantify interobserver agreement.

Kappa values interpreted as:

<0: Poor

0–0.20: Slight

0.21–0.40: Fair

0.41–0.60: Moderate

0.61–0.80: Substantial

0.81–1.00: Almost perfect

The study demonstrated substantial interobserver agreement, supporting reproducibility of the MsPath scoring system.

All statistical analyses were performed using SPSS (version XX), and a p-value of <0.05 was considered statistically significant.

## CHAPTER 03

### Discussion and conclusion

#### 3.1 Prediction of MSI-H CRC

This study confirms that MsPath score, combined with histological features, is a reliable predictor of MSI-H colorectal cancers.

High MsPath scores were strongly associated with MMR-deficient tumours (p < 0.05).

The independent predictors identified in logistic regression (high MsPath score, mucinous differentiation, lymphocytic infiltration) align with previous studies describing clinicopathological hallmarks of MSI-H tumours, particularly right-sided colorectal cancers.

This supports the clinical utility of MsPath scoring in triaging cases for further molecular testing, potentially saving resources in low-prevalence settings.

#### 3.2 Interobserver Reproducibility

Substantial agreement between observers indicates that MsPath scoring can be consistently applied in practice.

Disagreements were largely due to borderline histological features or technical issues, suggesting that additional training or standardized guidelines could further improve reliability.

#### 3.3 Limitations

The sample size of cases that underwent immunohistochemistry (IHC) for mismatch repair (MMR) proteins was relatively small, with only 46 colorectal carcinoma cases included. This limited number may have reduced the statistical power of the study and restricts the generalisability of the findings to the broader colorectal cancer population.

Selection bias is another important limitation, as only cases selected for MMR IHC were included in the analysis. These cases may not fully represent the entire spectrum of colorectal carcinomas encountered in routine diagnostic practice, particularly with respect to tumour stage, histological subtype, and anatomical distribution.

Technical variability represents an important limitation of this study. At the time the study was conducted, immunohistochemistry (IHC) for mismatch repair (MMR) proteins was not routinely established in the laboratory, and medical laboratory technologists (MLTs) had limited prior experience with these specific immunostains. Consequently, a local staining protocol had to be developed and optimised. This process involved multiple trial runs with adjustments to antigen retrieval methods, antibody dilutions, incubation times, detection systems, and the use of different reagent batches during the pre-assessment phase. Such repeated optimisation may have introduced variability in staining quality and intensity, which could have influenced both MsPath scoring and the interpretation of MMR protein expression.

In addition, the optimisation and validation runs consumed a proportion of the available tissue and antigenic material, thereby limiting the number of cases that could ultimately be included for definitive MMR IHC analysis.

Furthermore, the optimisation and validation process consumed not only tissue and antigenic material but also other essential laboratory consumables, including glass slides, coverslips, and reagents required for antibody dilutions and detection systems. In addition, limitations in laboratory staff availability to prepare and process immunohistochemical slides contributed to constraints on throughput. These combined resource and logistical factors restricted the number of cases that could be processed and included in the final analysis.

Also differences in tissue fixation, processing, section thickness, and interobserver interpretational variability may have affected the assessment of both histomorphological features included in the MsPath score and the evaluation of IHC results. Although standard reporting criteria were followed, subtle variations in staining quality and nuclear expression could have influenced classification of MMR status, particularly in borderline or heterogeneous cases.

Finally, this was a single-centre study conducted in a specific laboratory setting with locally optimised protocols. Therefore, external validation in larger, multi-centre cohorts with standardised and well-established MMR IHC protocols is required to confirm the reproducibility and broader applicability of these findings.

#### 3.4 Conclusion

Right-sided colorectal carcinomas in this Sri Lankan cohort more frequently exhibited histological features suggestive of MSI-H, including mucinous differentiation, TILs, and Crohn-like lymphoid reaction. MsPath scoring correlated with MMR deficiency in the IHC-tested subset, but its predictive value is limited without immunohistochemical confirmation. IHC using a two-antibody panel (PMS2 and MSH6) proved to be a feasible, cost-effective, and reliable method for MSI screening in resource-limited settings.

#### 3.5 Recommendations

Use MsPath score in conjunction with histological features to prioritize cases for MSI testing, especially in resource-limited settings.

Implement standardized guidelines and training for MsPath scoring to reduce interobserver variability.

Increase sample size and perform multicentre studies to validate these predictors of MSI-H status.

Consider molecular confirmation of MSI status in cases with borderline MsPath scores or discordant IHC findings.

Explore integration of MsPath scoring with additional biomarkers (e.g., BRAF mutation, MLH1 promoter methylation) for enhanced predictive accuracy.

## Data Availability

All data produced in the present work are contained in the manuscript

## Annexures

## Annexure-01 – Photomicrographs of a few cases from the study

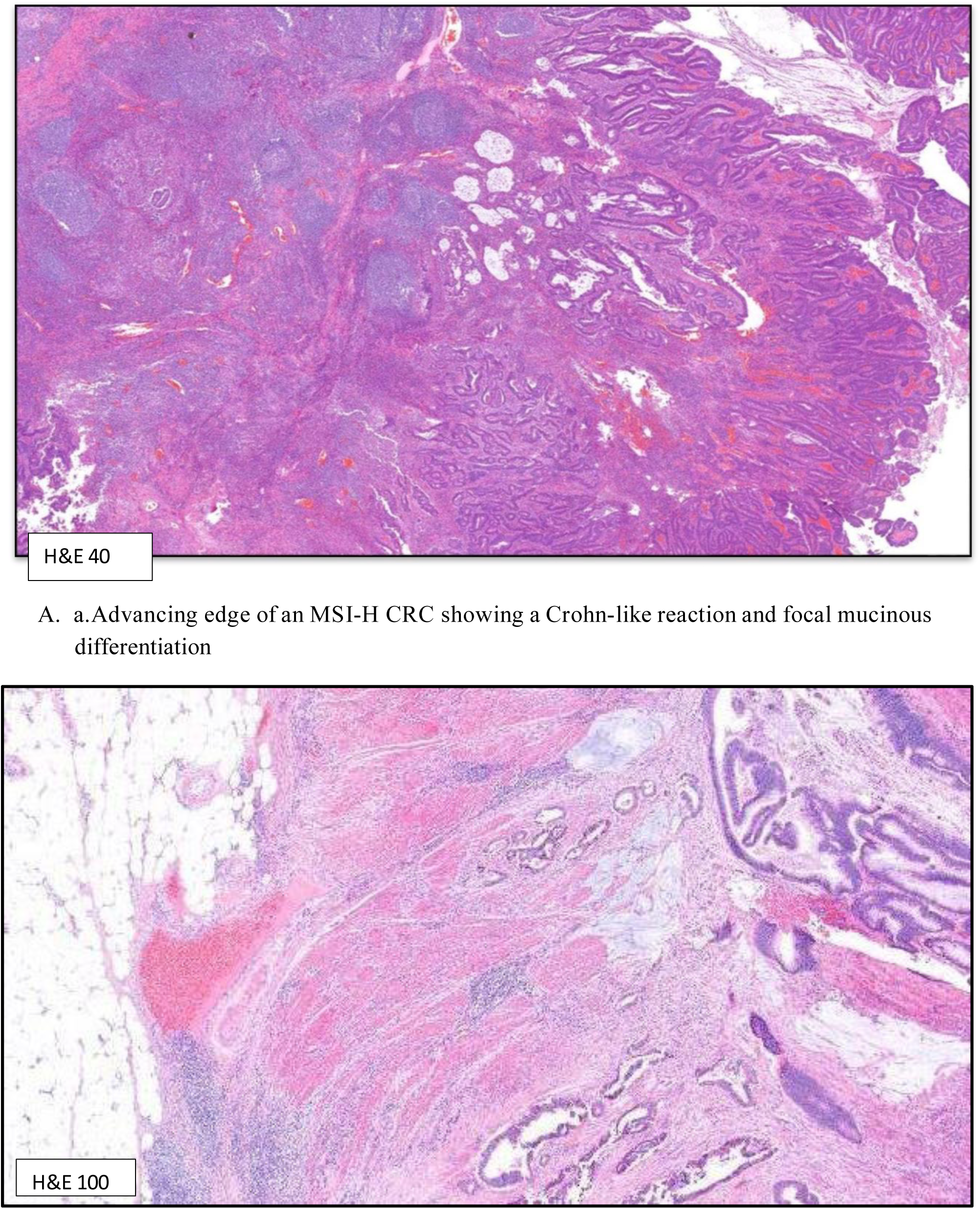
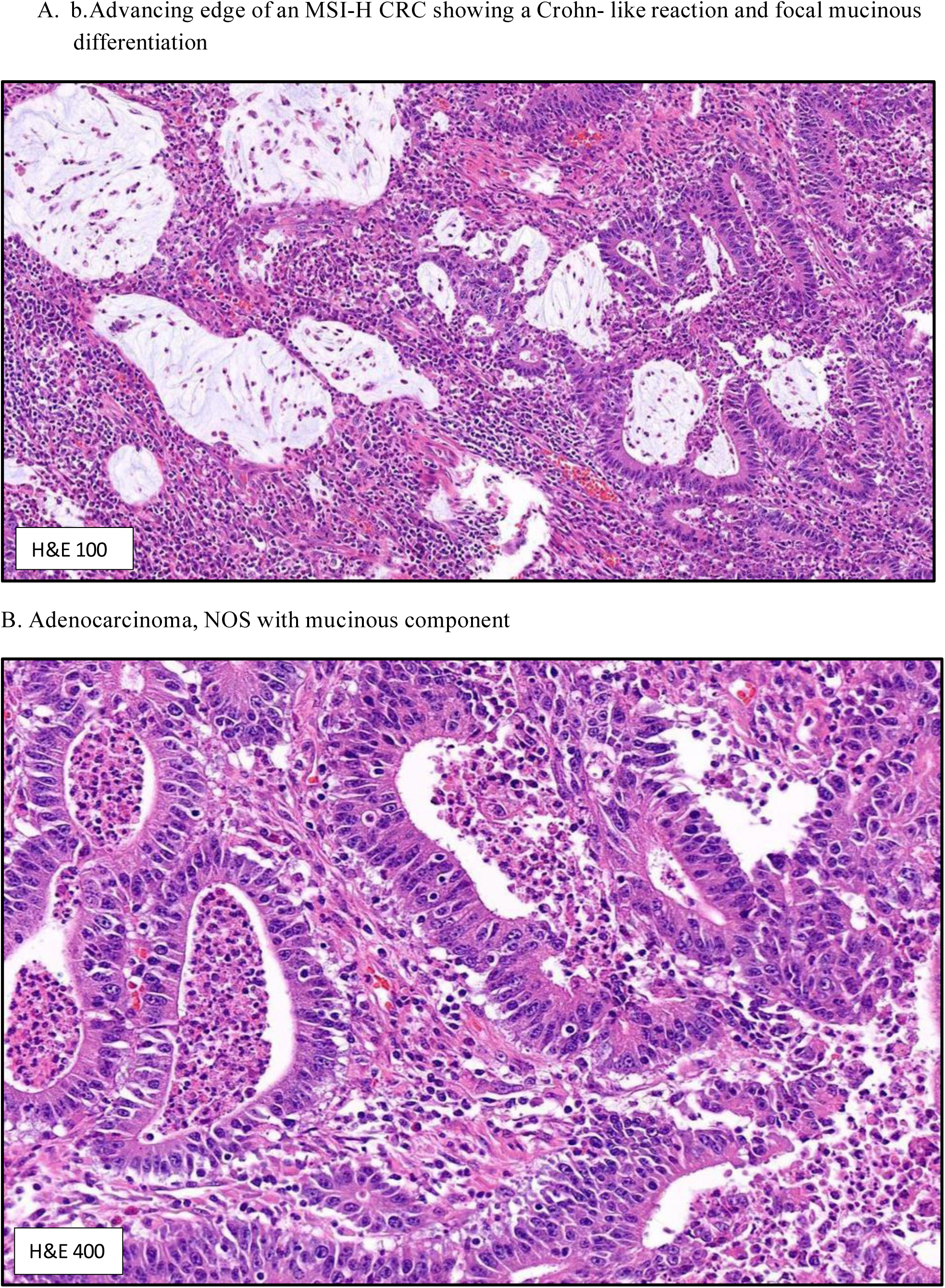
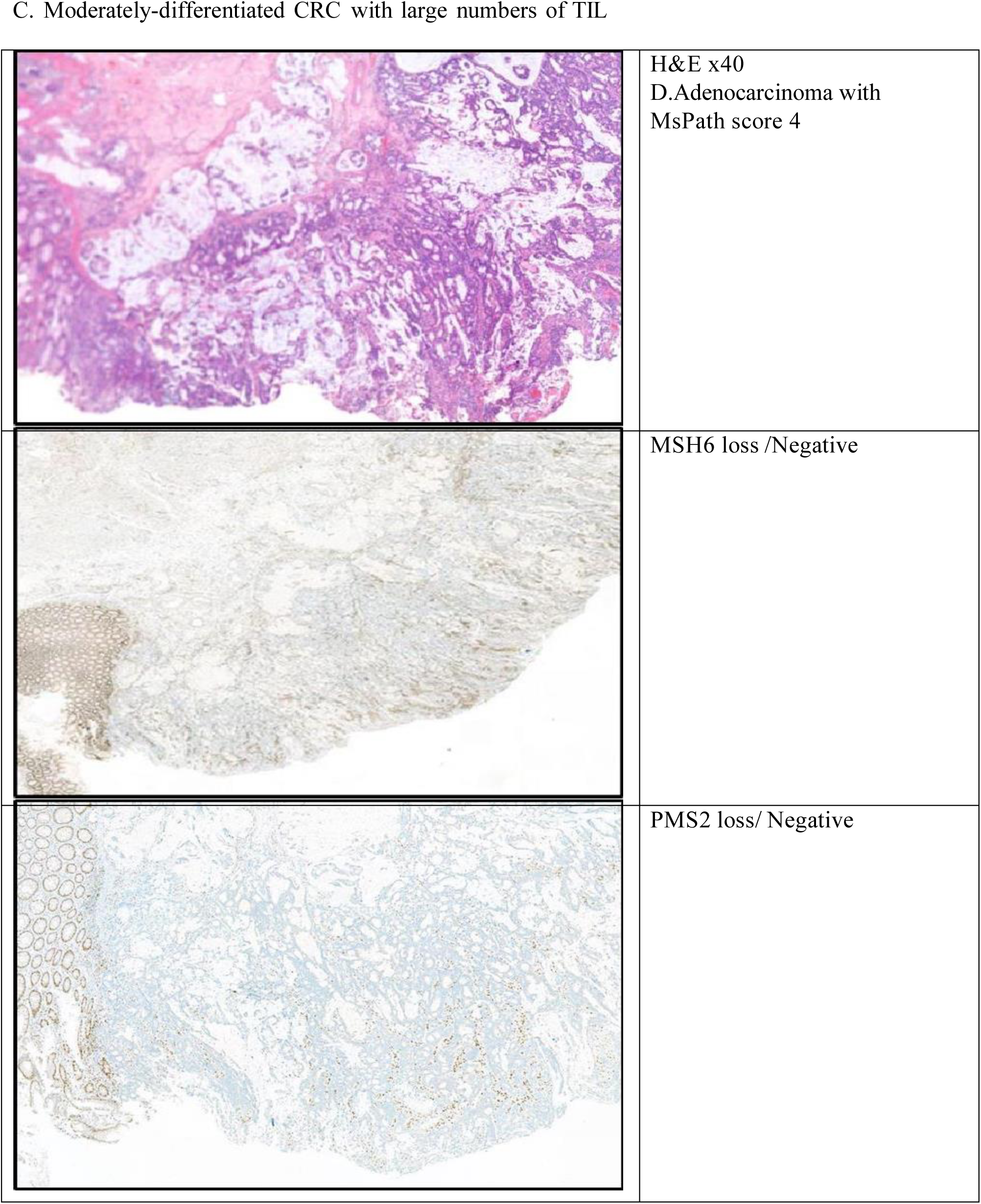
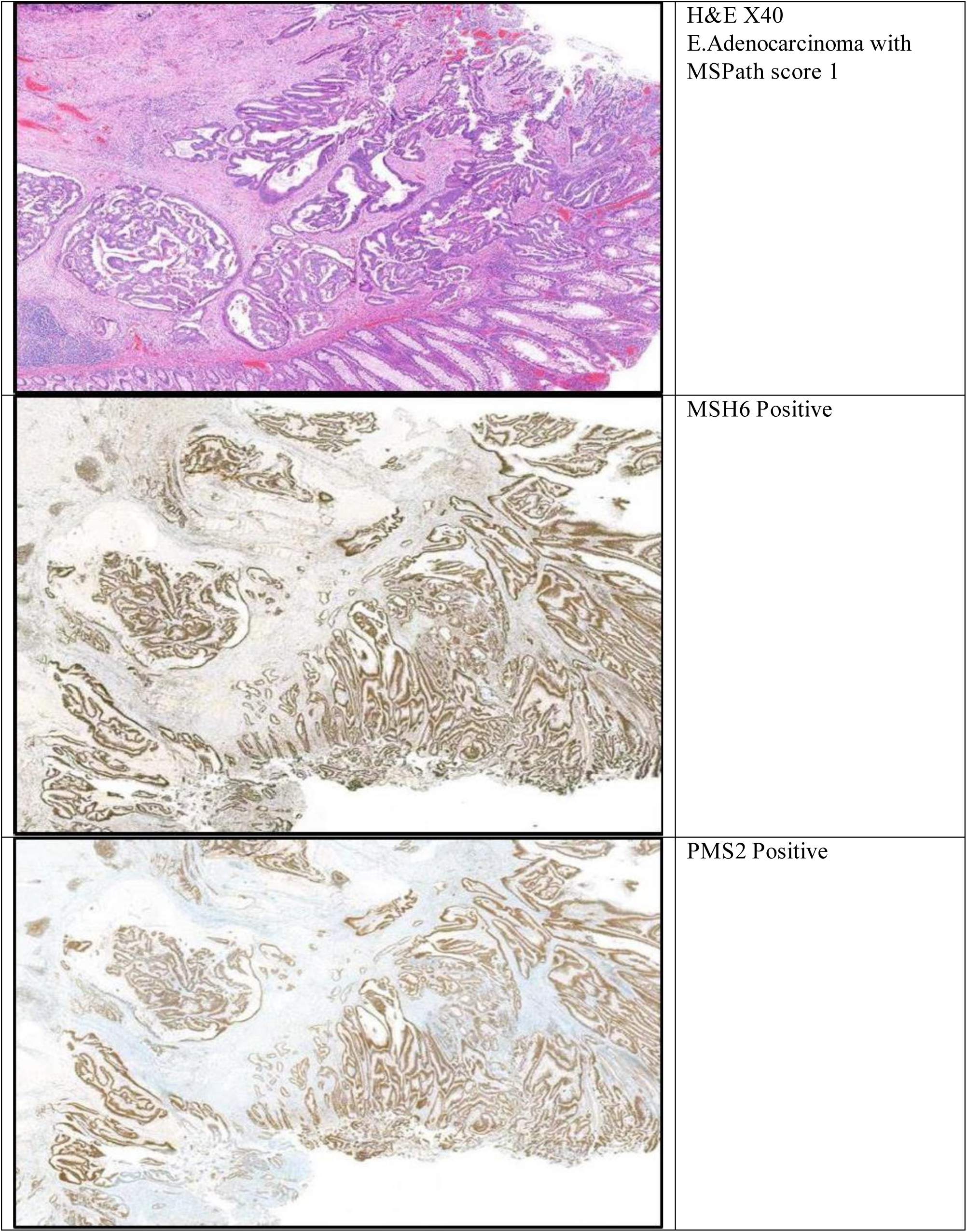

## Annexure II Data collection sheet

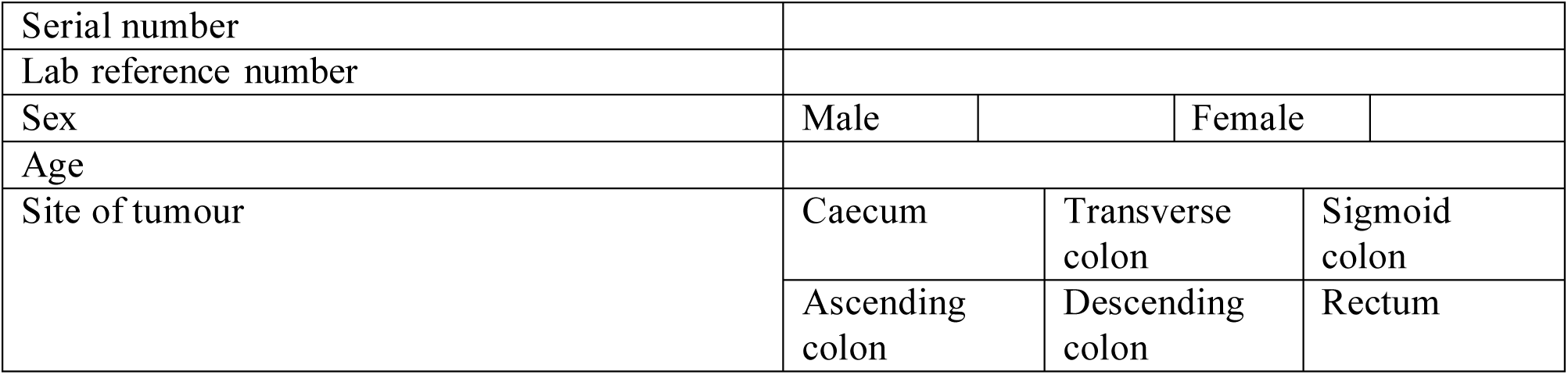

Assessment of the histological features according to the definitions (Present/Absent)

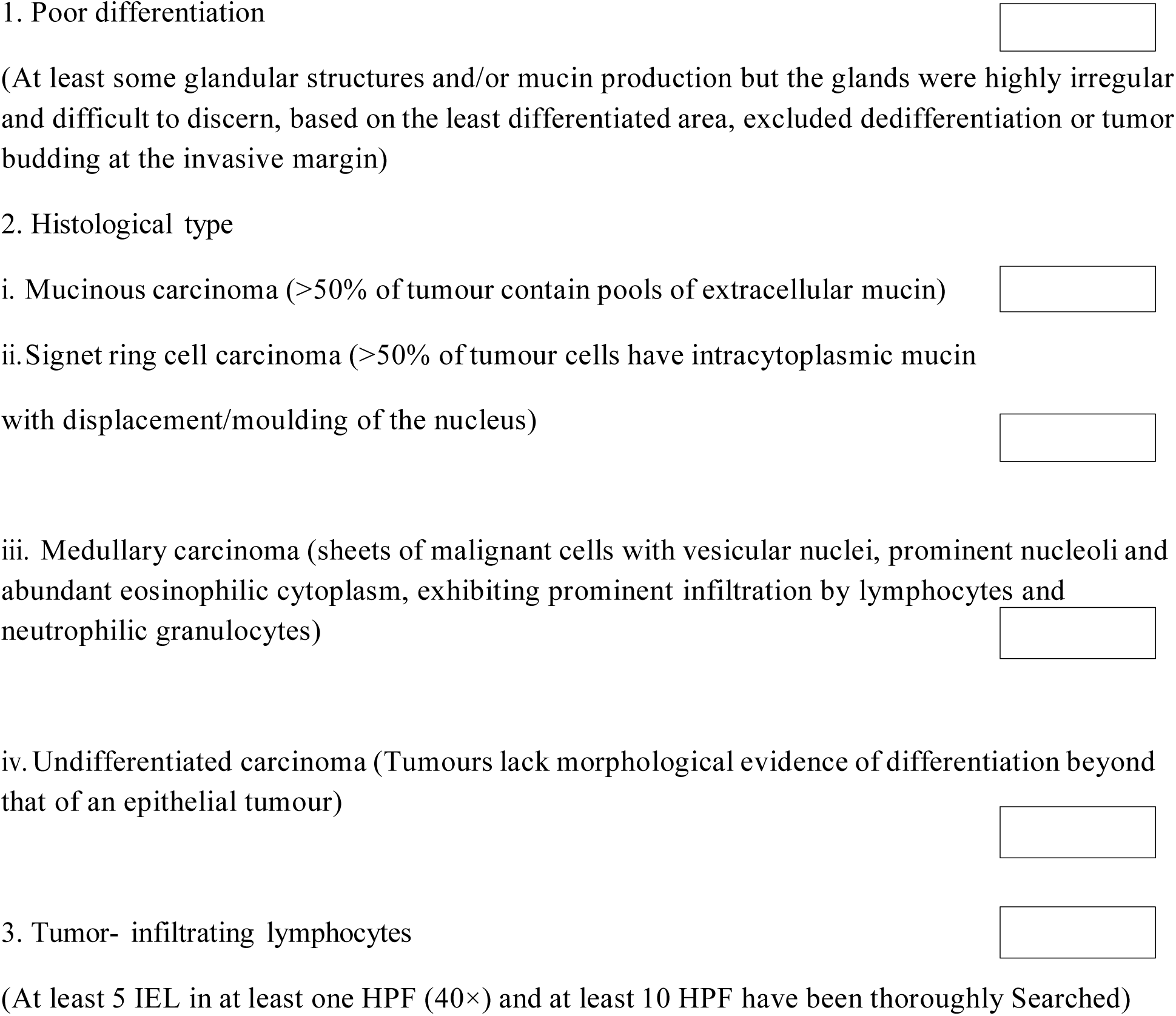
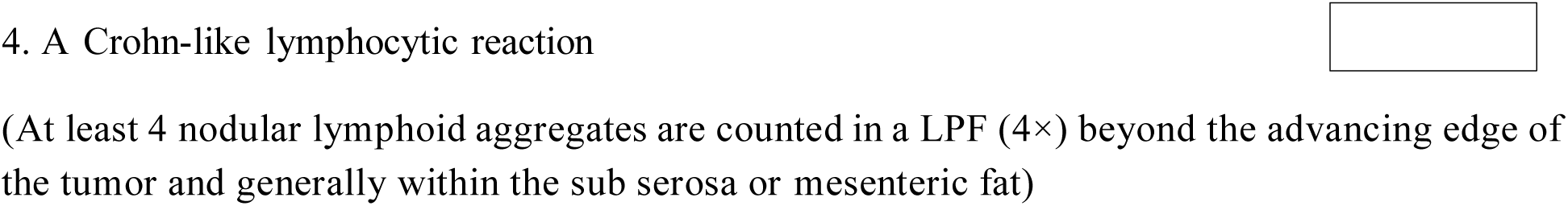

To calculate the MsPath Score

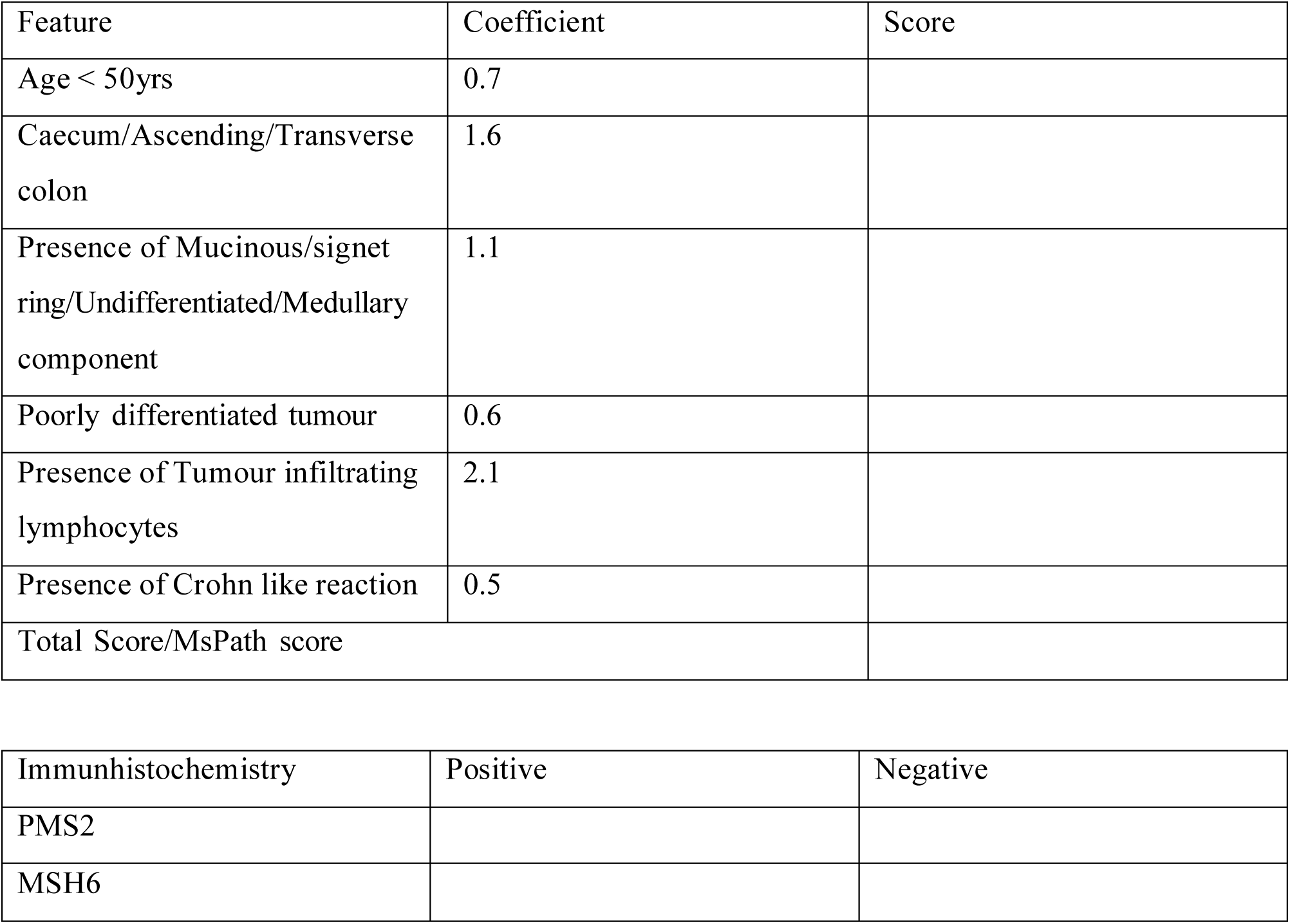

| Feature | Coefficient | Score |
| --- | --- | --- |
| Age < 50yrs | 0.7 |  |
| Caecum/Ascending/Transverse colon | 1.6 |  |
| Presence of Mucinous/signet ring/Undifferentiated/Medullary component | 1.1 |  |
| Poorly differentiated tumour | 0.6 |  |
| Presence of Tumour infiltrating lymphocytes | 2.1 |  |
| Presence of Crohn like reaction | 0.5 |  |
| Total Score/MsPath score |  |  |

| Immunohistochemistry | Positive | Negative |
| --- | --- | --- |
| PMS2 |  |  |
| MSH6 |  |  |

## Annexure III

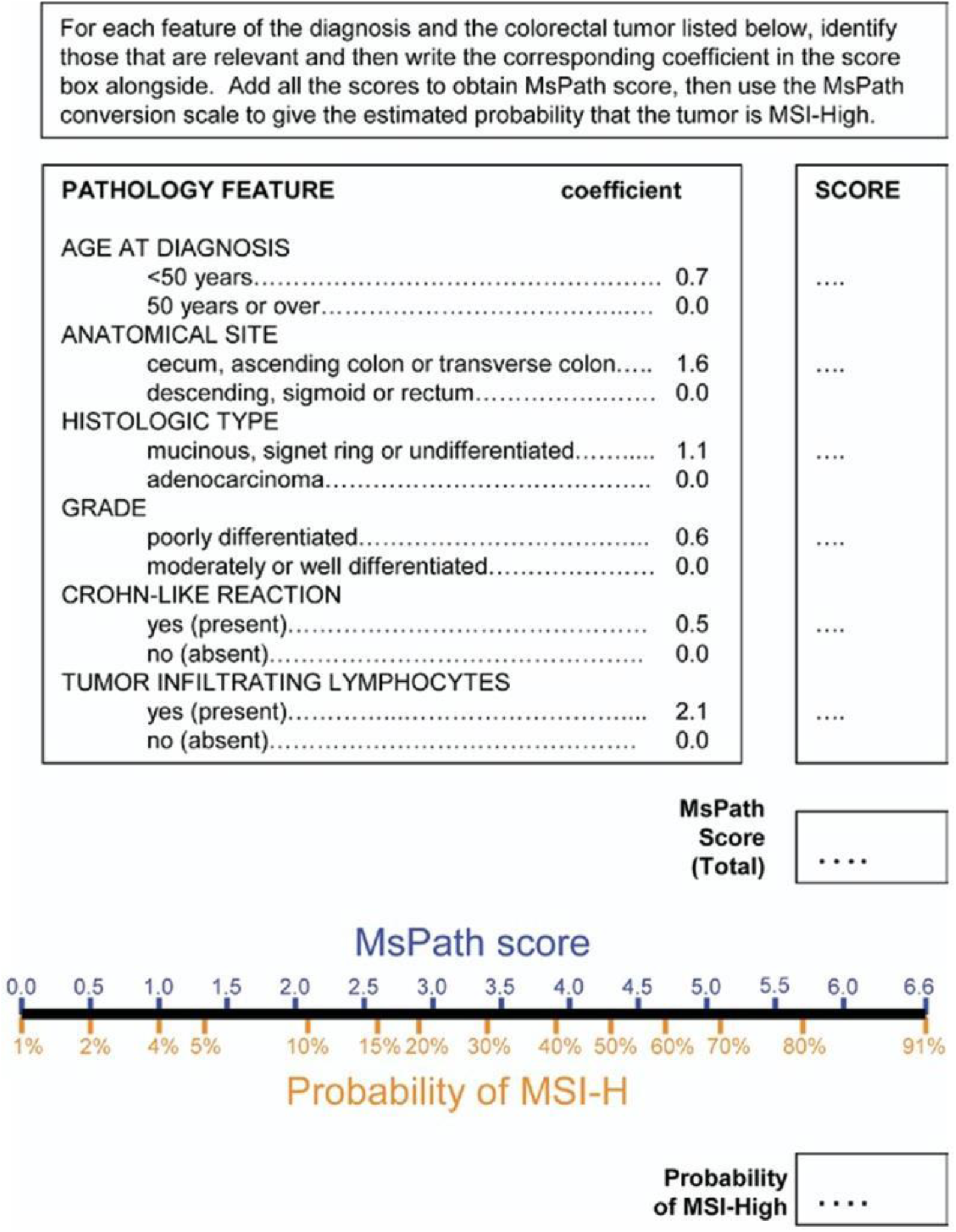

## Annexure-IV Immunohistochemical Staining Procedure

- Immunohistochemical staining was performed with on-slide control tissue, with both case and control tissue mounted on the same slide.
- Liquid-form primary antibodies were used for staining.
- Multiple antibody dilution series were tested on positive control slides to determine the optimal working dilutions for MSH6 and PMS2.
- Final antibody dilutions selected were:
- PMS2: 1:50 (antibody: antibody diluent)
- MSH6: 1:100 (antibody: antibody diluent)
- Paraffin wax blocks containing unfixed tissue were excluded from the study.
- Staining was carried out using a fully automated immunohistochemistry analyzer (BioGenex XMatrix Lite).
- An indirect immunoassay technique was employed.
- Background staining was minimized by increasing the number of washing steps.
- Washing steps were considered essential for achieving optimal immunohistochemical staining quality.
- Phosphate-buffered saline (PBS, pH 7.4) containing a surfactant was used as the wash buffer.
- The wash buffer facilitated uniform distribution of antibodies and reagents, reducing non-specific staining and ensuring consistent results.

## Annexure V Approval letter by board of Study in Pathology

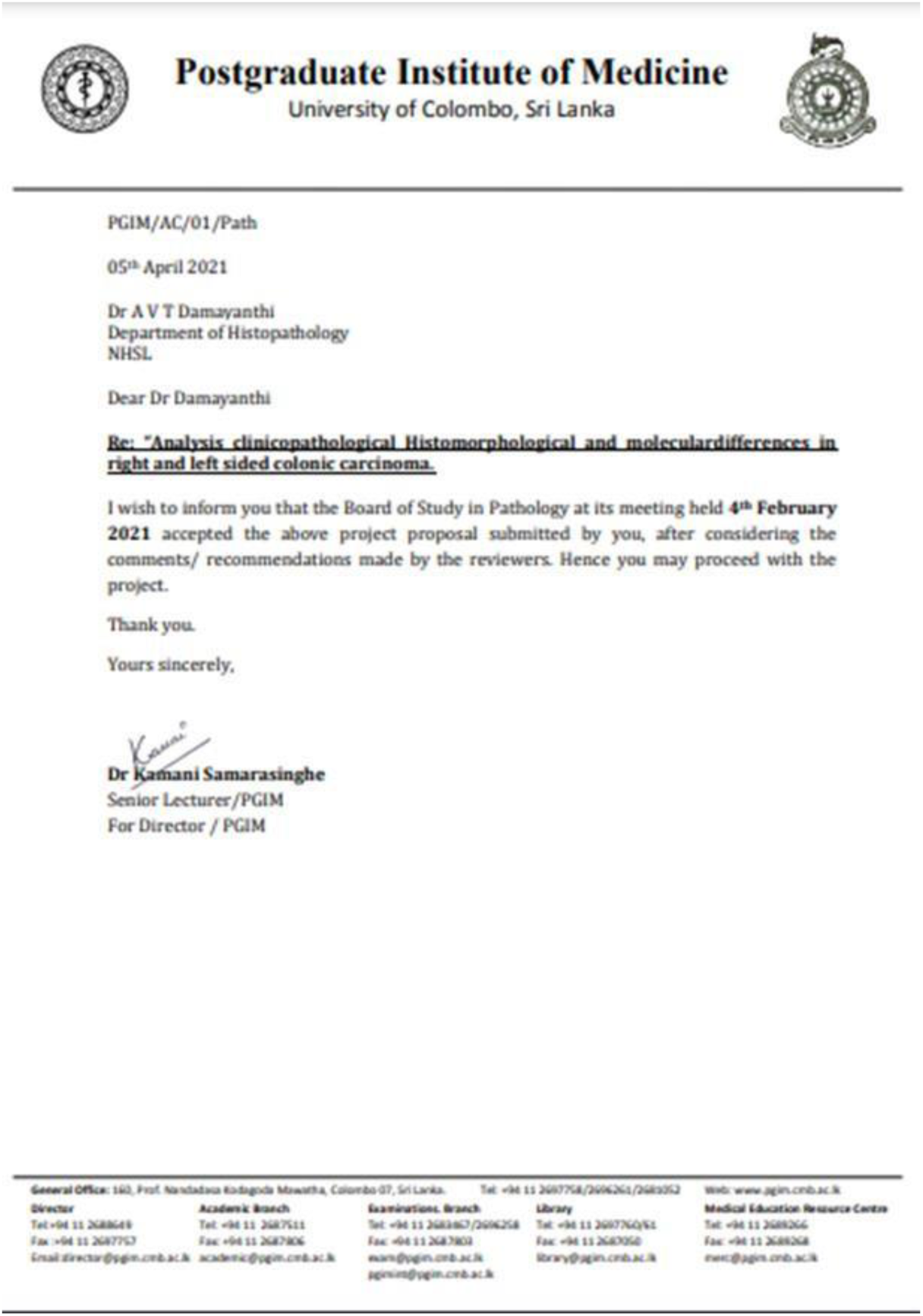

## Annexure-VI Ethical approval letter received by the Research Ethical review committee of the National Hospital of Sri Lanka

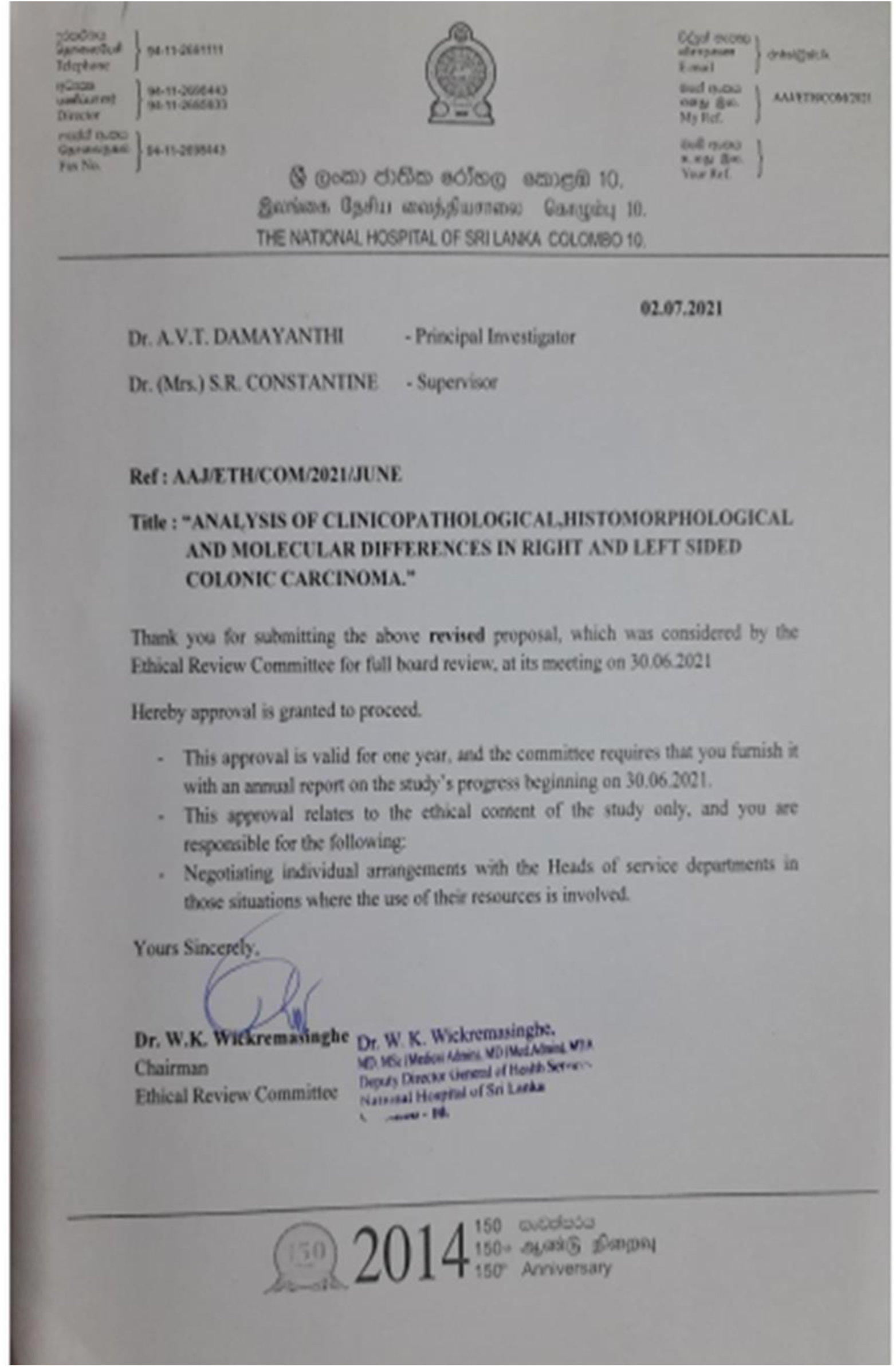

## Annexure VII Budget estimation

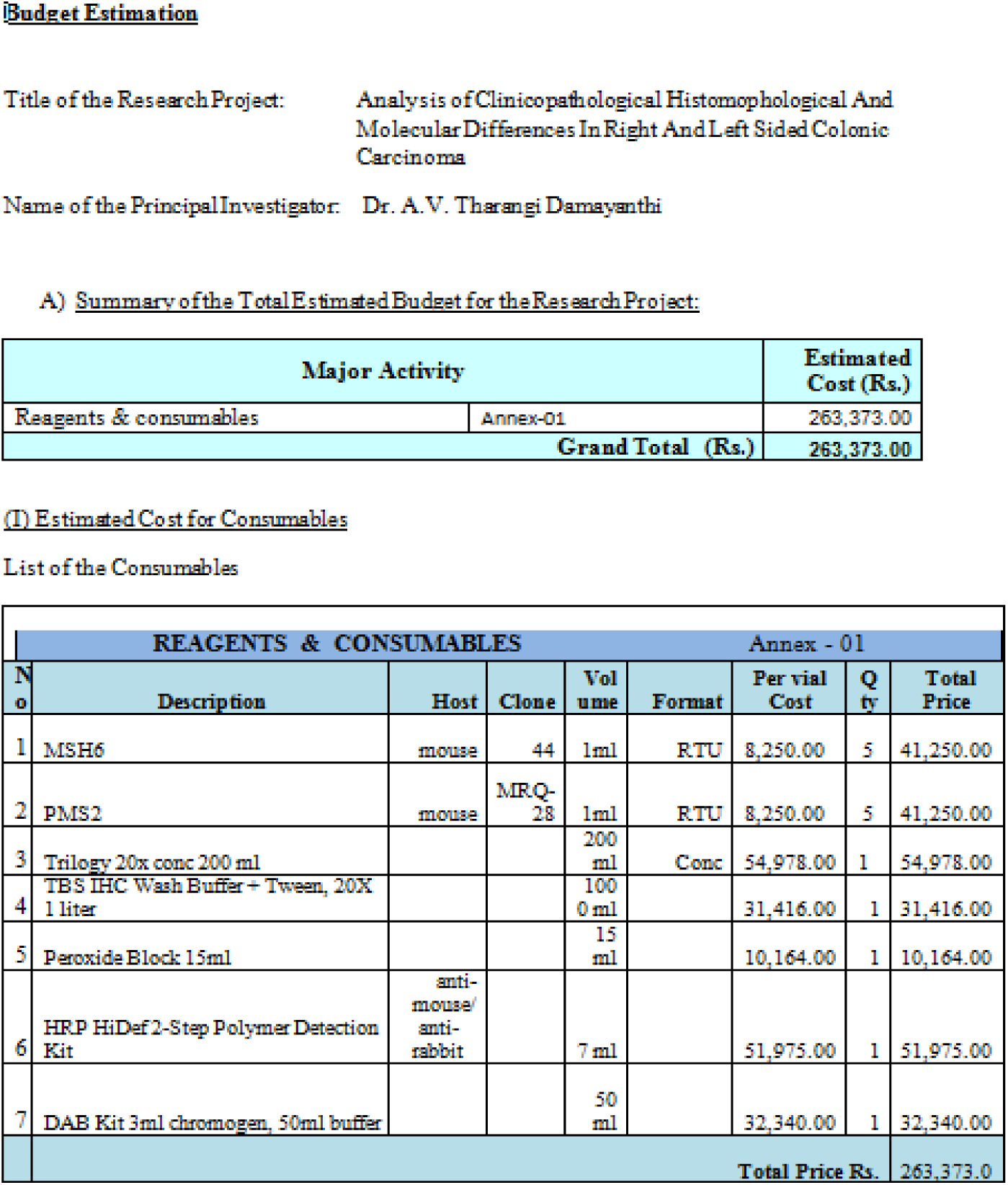

